# Long-term effects of increased adoption of artemisinin combination therapies in Burkina Faso

**DOI:** 10.1101/2021.08.20.21262380

**Authors:** Robert J. Zupko, Tran Dang Nguyen, Anyirékun Fabrice Somé, Thu Nguyen-Anh Tran, Jaline Gerardin, Patrick Dudas, Dang Duy Hoang Giang, Kien Trung Tran, Amy Wesolowski, Jean-Bosco Ouédraogo, Maciej F. Boni

## Abstract

Artemisinin combination therapies (ACTs) are the WHO-recommended first-line therapies for uncomplicated *Plasmodium falciparum* malaria. The emergence and spread of artemisinin-resistant genotypes is a major global public health concern due to the increased rate of treatment failures that result. This is particularly germane for WHO designated ‘high burden to high impact’ (HBHI) countries, such as Burkina Faso, where there is increased emphasis on improving guidance, strategy, and coordination of local malaria response in an effort to reduce the prevalence of *P. falciparum* malaria. To explore how the increased adoption of ACTs may affect the HBHI malaria setting of Burkina Faso, we added spatial structure to a validated individual-based stochastic model of *P. falciparum* transmission and evaluated the long-term effects of increased ACT use. We explored how *de novo* emergence of artemisinin-resistant genotypes, such as *pfkelch13* 580Y, may occur under scenarios in which private-market drugs are eliminated or multiple first-line therapies (MFT) are deployed. We found that elimination of private market drugs would result in lower treatment failures rates (between 11.98% and 12.90%) when compared to the status quo (13.11%). However, scenarios incorporating MFT with equal deployment of artemether-lumefantrine (AL) and dihydroartemisinin-piperaquine (DHA-PPQ) may accelerate near-term drug resistance (580Y frequency ranging between 0.62 to 0.84 in model year 2038) and treatment failure rates (26.69% to 34.00% in 2038), due to early failure and substantially reduced treatment efficacy resulting from piperaquine-resistant genotypes. A rebalanced MFT approach (90% AL, 10% DHA-PPQ) results in approximately equal long-term outcomes to using AL alone but may be difficult to implement in practice.

## Introduction

Despite the United Nations’ Millennium Development Goals leading to substantial reductions in the global burden of infectious disease, malaria remains a serious public health risk in many parts of Africa, Asia, and South America. *Plasmodium falciparum* malaria remains holoendemic in at least eleven countries, mainly in western and central Africa, and these countries have recently come under a WHO grouping called high burden to high impact (HBHI) which recognizes that the most severely affected malaria-endemic countries will require additional resources and greater policy imagination [1]. The two primary concerns in HBHI countries are high and widespread vectorial capacity and insufficient access to high-efficacy antimalarial therapies. Traditionally, drug-resistance concerns in holoendemic settings are of secondary importance because of the historical relationship between low-transmission regions and drug-resistance emergence and the relatively weak selection pressure imposed by lower levels of antimalarial use.

Artemisinin-resistant phenotypes were first identified in 2007-2008 in western Cambodia and have since been linked to a number of point mutations in the *P. falciparum kelch13* gene [2,3]. Since the identification of these molecular markers, artemisinin resistance has been found in in Bangladesh, Guyana, Laos, Myanmar, Papua-New Guinea, Thailand, Rwanda, and Vietnam [1,3–8]. Artemisinin-resistant parasites can either be imported from other regions (e.g., spread across Southeast Asia), or they may emerge independently as in Guyana, Papua-New Guinea, and Rwanda [3,4,7]. This raises the possibility that HBHI countries may be at risk for artemisinin resistance appearing due to *de novo* emergence or the importation of a resistant parasite.

Burkina Faso has been identified as an HBHI nation and malaria remains the leading cause of hospitalization in the general population (45.8% of all hospitalizations) as well as for children under five (48.2%) [1,9]. Malaria transmission is endemic throughout the country and seasonal transmission is aligned with the rainfall patterns of the three seasonal zones. The northern Sahelian region has a short-peak transmission season of approximately three months, increasing to four months in the central Sudano-Sahelian region, and reaching about five months in the southwestern Sudanian region [10]. Artemisinin combination therapies have been in use in Burkina Faso since their adoption in 2005, resulting in significant reductions in malaria prevalence [1,10,11]. Alongside increasing access to ACTs, local public health officials have also credited broader adoption of insecticide-treated mosquito nets (ITNs), higher levels of indoor residual spraying (IRS), and new programs in intermittent preventive treatment of pregnant women (IPTp) and seasonal malaria chemoprevention (SMC) [12]. The national goal for SMC is that it be provided to all children aged 3-59 months by 2020 [12].

Presently, there are no signs of artemisinin resistance in Burkina Faso, as no molecular markers have been observed. However, recent studies conducted as part of WHO recommended periodic efficacy surveillance efforts, along with other clinical drug trials, suggest that first-line ACT therapies may have reduced efficacy in children aged 6-59 months [13], and detectable parasitemia 72 hours after treatment with artemether–lumefantrine (AL) has been observed [14]. While not as pressing a concern as clear genomic markers for drug resistance, these findings do suggest that alterations to the present drug policy may be needed, as noted by Gansané et al. [13].

The widespread adoption of ACTs has led to reductions in malaria prevalence in the past 10 years, but the widespread use of ACTs within Burkina Faso also places selective pressure on the parasite to evolve resistance. The impact that artemisinin-resistant *P. falciparum* would have in Burkina Faso is currently unknown; however, ensuring the continued efficacy of ACTs is a top priority for all national malaria control programs in Africa. Accordingly, the development of spatial, individual-based stochastic models allows for projections to be made as to when and where artemisinin resistant parasites may appear and how drug policies may affect the evolution of drug resistance in the parasite. In this study we have calibrated a spatial individual-based model for Burkina Faso. This model has been used to explore two broad concerns: the possible emergence or importation of artemisinin resistance under the current health care regime and the impact that various drug policy interventions may have on artemisinin resistance.

## Methods

### Model Design

This study builds upon the individual-based stochastic model of malaria transmission previously developed by Nguyen et al. [15] to incorporate space and geography. The model was parametrized to fit the annual mean Malaria Atlas Project *Pf*PR_2-10_ projections for Burkina Faso in 2017 on a 5km-by-5km cell (or pixel) basis with a total of 10,936 pixels modeled (approximately 273,400 sq.km) [11]. Individual movement was fit to available travel survey data for both the destination and frequency of travel (Fig. 1d) [16–18]. In the individual-based model, individuals have attributes relevant to the spread and individual response to a *P. falciparum* infection, such as age, attractiveness to mosquitos, number of parasite infections, genotypes of infecting parasites, level of parasitemia, level of immunity, and current drug concentrations during and after treatment. In addition to the new of spatial components, the individual infection model was revised to incorporate a more realistic sporozoite challenge (S1 File §5.1). The spatial component of the model allows for variables such as climate, heterogeneous access to treatment, and individual movement to be incorporated into the simulation.

**Fig. 1:**
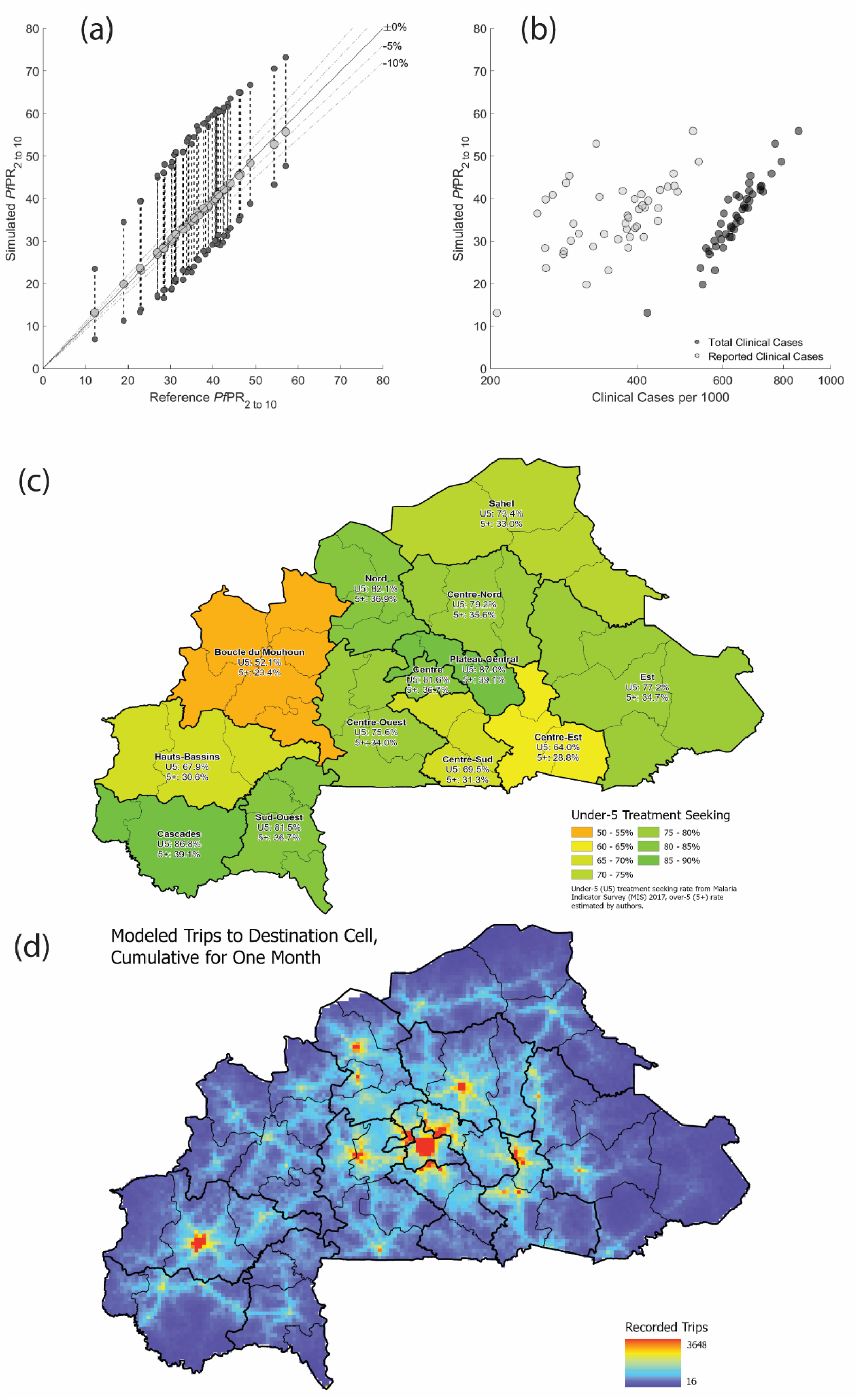
Model calibration and validation. (**a**) The first point of validation for the model is the comparison of the simulated *Pf*PR_2-10_ values versus the reference *Pf*PR_2-10_ values from the Malaria Atlas Project (Weiss et al. 2019). Each light gray dot corresponds to one province of Burkina Faso. The annual mean of the simulated *Pf*PR_2-10_ values (light gray dots) are plotted along the y-axis while the reference *Pf*PR_2 to 10_ is plotted along the x-axis. The upper and lower dark gray dots indicate the annual peak and minimum of the seasonal *Pf*PR_2-10_ for the given province. **(b)** Clinical (i.e., symptomatic) cases of malaria plotted against the *Pf*PR_2-10_, for every province. Black dots correspond to every clinical case in the simulation, while gray dots are the clinical cases that would be reported (according to drug coverage numbers). (**c**) The treatment seeking rate used to inform the model is dependent upon the region of Burkina Faso that an individual is in, with the under-5 treatment rate ranging from a low of 52.1% to a high of 87.0%. Note that the over-5 treatment seeking rate is adjusted to be 45% of the under-5 treatment seeking rate to account for the lower treatment seeking rate in older demographic groups. (**d**) A second point of validation for the model is ensuring that movement is consistent with expected patterns. The heatmap shows the number of trips to the destination cell over the course of a single month in the model. Note that the distribution indicates that while there is always a low chance that an individual may visit any cell, individuals are primarily attracted to major population centers. Subfigure (c) reproduced from Zupko et al. [18], Burkina Faso administrative boundaries from the World Bank Group [19].

In order to calibrate the model for Burkina Faso, we started by first preparing multiple geographic information system (GIS) raster files for Burkina Faso with a scale of 5km pixels, or adjusted rasters through aggregation to conform to the 5km scale when necessary. These rasters in turn contain such information as the treatment coverage, climate zone, and the population size. Upon model initialization, there is a population of approximately 3.6 million individuals or about 25% of the 2007 population of Burkina Faso, who are distributed based upon the population density of the country [20]. This population is allowed to grow and move throughout the simulated landscape, carrying any *P. falciparum* populations that they may be infected with. Data are collected from the model starting in the simulated year 2018 which approximates the most recent Malaria Indicator Survey for Burkina Faso used for model calibration [21]. Simulation runs using 100% versus 25% of the population size are not found to be qualitatively different.

When individuals in the model are infected, the asexual parasite density is modeled and the levels of parasitemia and immune response are used to determine symptoms occurrence and treatment seeking behavior. Depending upon the individual and their immune response, they may progress towards clinical symptoms or an asymptomatic infection. When presenting with symptoms, individuals in the model seek treatment at different rates based on their age group: under-5 treatment rates were based upon the Malaria Indicator Survey rates [21], while the over-5 treatment rates were adjusted to be 55% lower (relative scale) than the under-5 treatment rate to account for lower rates of treatment seeking among adults [22]. Both treatment rates are increased by 3% each model year to account for the expected increase in access to treatment over time [23]. Therapies given to individuals use a simplified single-compartment pharmacokinetic/pharmacodynamic (PK/PD) model with PK/PD parameters calibrated to reflect the expected efficacies of specific antimalarial compounds on particular *P. falciparum* genotypes [24], and parasitemia levels after 28 days are used to determine treatment failure.

The primary measure for artemisinin resistance in the model is the frequency of the 580Y mutation in *pfkelch13*. Upon model initialization the frequency of 580Y is zero and upon reaching the model calibration time point, the locus is allowed to mutate (with a previously calibrated probability [25]) once per day in the presence of drugs (Fig. 2). The simulation focuses on mutations in the presence of drugs due to the fitness penalty that is associated with most drug resistance markers [26–28], effectively resulting in drug-resistant genotypes being selected against in the absence of drug pressure. The *de novo* emergence rate of 580Y alleles – an unknown value in general contexts – was set using a previous model alignment exercise in an environment with 10% *Pf*PR_2-10_ and 40% drug coverage with DHA-PPQ only [25]; under this alignment 580Y alleles progress from 0.00 to 0.01 allele frequency in seven years. Accordingly, the progression from 0.00 to 0.01 allele frequency in the presence of the drug mixture in Burkina Faso may be faster (or slower). Other genetic makers associated with drug resistance (*plasmepsin-2,3* double copy, *pfmdr1* double copy, *pfcrt-K76, pfmdr1-N86, pfmdr1-184F*) are included in the model and operate in a similar fashion to 580Y with the model initialized with the sensitive genotype. For each scenario, a total of 50 replicates are performed to ensure sufficient coverage of outlier simulation results.

**Fig. 2:**
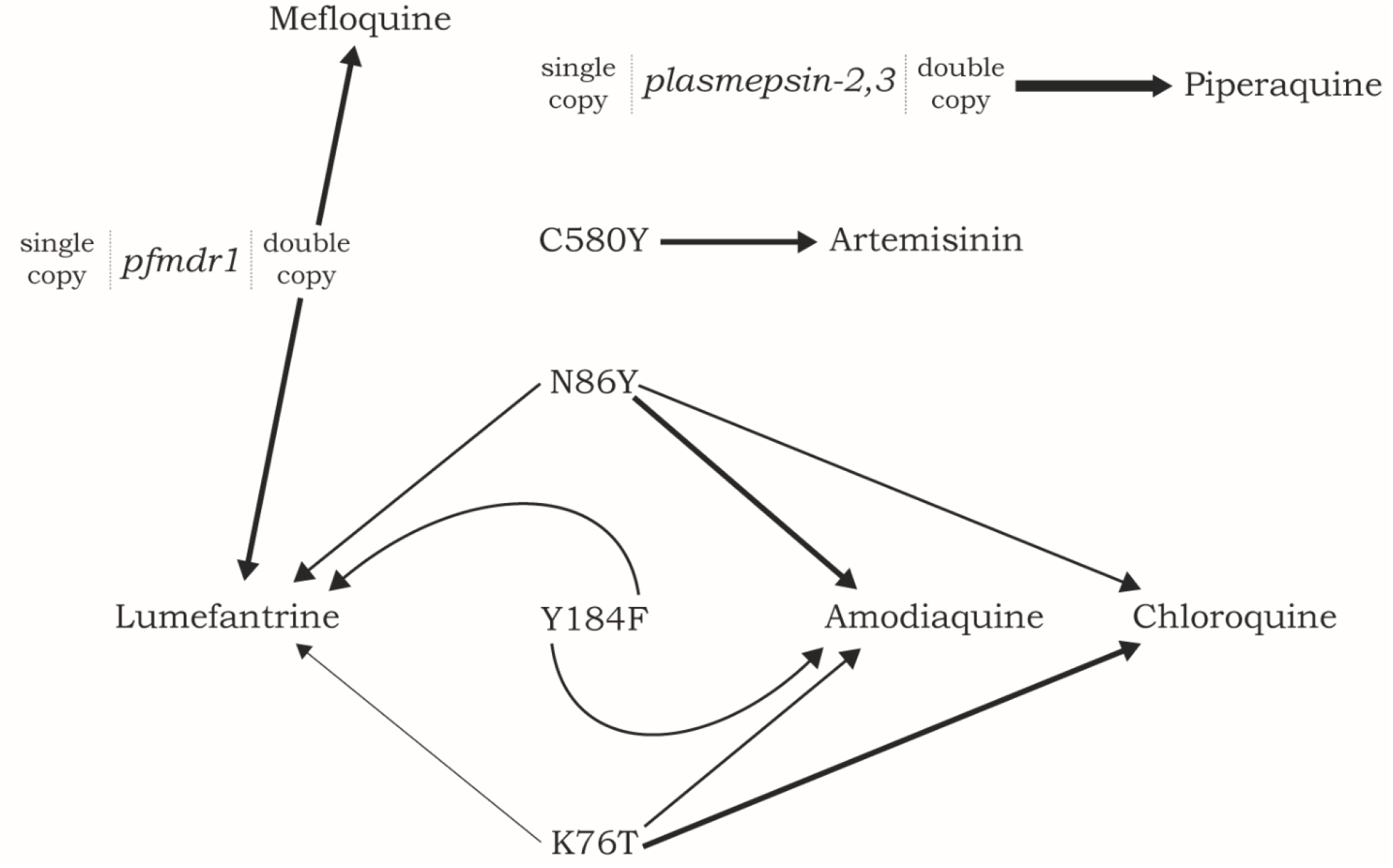
Diagram showing pleiotropy of loci affecting drug-resistance phenotypes. An arrow pulling on an allele or genotype indicates that that drug selects for that allele or copy-number variant. For example, amodiaquine selects for alleles 86Y and Y184 in the *pfmdr1* gene and allele 76T in the *pfcrt* gene. Thicker arrows indicate stronger selective pressure according to the drug-by-genotype interactions developed by Nguyen et al. [24].

To evaluate the impact that various drug policies have on treatment, the simulation tracks the number of clinical cases (i.e., individuals with clinical symptoms), the reported number of cases (i.e., individuals with clinical symptoms receiving medical care), treatment failures (i.e., individuals with clinical symptoms for whom the prescribed treatment was not effective through day 28), and the number of individuals with clinical symptoms who did not seek treatment. Additionally, the genotype of the parasite, or parasites in the case of a multiclonal infection, is tracked across all individuals in the model.

### Model Validation

The spatial model of Burkina Faso has pixels with a population of at least one individual per pixel at model initialization, allowing for comparisons to the annual mean *Pf*PR_2-10_ estimates for year 2017, using the first data set released by the Malaria Atlas Project [11]. Each pixel is assigned a transmission parameter *β* which corresponds, approximately, to the local force of infection or vectorial capacity at that location. Individual pixel *Pf*PRs were calibrated by finding a *β* that results in the appropriate annual mean *Pf*PR_2-10_ given a cell’s population size, treatment coverage, seasonal transmission pattern, and reference *Pf*PR_2-10_. The predicted population-weighted annual mean *Pf*PR_2-10_ values for each cell were then aggregated to the province level for comparison to the Malaria Atlas Project values (Fig. 1a). Individual movement was added into the model by using a modified gravity model [18] that was re-fit from data presented in Marshall et al. [16,17]. Pixel-level and province-level *Pf*PR_2-10_ values were evaluated to ensure that movement did not have too large of an overall effect on *Pf*PR_2-10_ trends. All individual movement presumes an eventual return to the original cell of the individual (i.e., round trips only), although the length of the trip is variable and may involve multiple stops. Province-level simulated *Pf*PR_2-10_ values, with the full model (including movement), are compared to reference *Pf*PR_2-10_ data in Fig. 1. All mosquito bites in the simulation are presumed to be infectious, and a bitten individual is infected with the parasite if they are unable to withstand the initial exposure to the sporozoite (S1 File §5.1).

### Scenarios Comparisons

All studies evaluated use a baseline business-as-usual configuration for Burkina Faso, in which the model is parameterized to match the Malaria Atlas Project prevalence in 2017 [11]. Drug coverage (i.e., the percentage of malaria-positive febrile individuals who are able to obtain drug treatment and choose to do so) for 2017 is set to be province-specific, based on data from a 2018 malaria indicator survey [21]. For symptomatic individuals that receive antimalarial treatment, the distribution of drugs used/prescribed is taken from this malaria indicator survey; however, since distribution is known only at a national level it is applied to the province-level drug coverage to get a final table of province-level treatment distributions. At a national level, the recommended first-line therapies are AL or dihydroartemisinin-piperaquine (DHA-PPQ), and artesunate (AS) or quinine for severe malaria. These co-exist with private-market treatments artesunate-mefloquine (ASMQ), amodiaquine (AQ), chloroquine (CQ), and sulfadoxine-pyrimethamine (SP) [21]. At a national level, 16.8% of individuals use antimalarials purchased in the private market and 83.2% receive an artemisinin combination therapy in the public-sector (91.6% AL, 6.6% ASAQ, 1.8% DHA-PPQ) for uncomplicated malaria in non-pregnant individuals. To account for the phase-out of ASAQ in 2018 as a former first-line therapy [12], the 6.6% usage was proportionally redistributed for the final business-as-usual public-sector treatment mix (93.3% AL, 6.7% DHA-PPQ). Access to treatment is assumed to go up every year in each province by about 3% starting with model year 2019 as the first year of improved access.

The distribution of treatments used in Burkina Faso suggests two possible drug policy interventions impacting the use of private market drugs and the deployment of nationally recommended first-line therapies. First, private-market drugs account for about 16.8% of treatments for febrile illness, with AQ monotherapy accounting for 73.8% of private-market treatments (12.4% of national treatments) [21]. Reduction or elimination of private-market purchases would ensure that individuals are more likely to receive a recommended first-line therapy (always an ACT) that has higher efficacy. Second, it may be possible to shift from AL as the primary single first-line therapy to the use of multiple first-line therapies (MFT) in which AL and DHA-PPQ are prescribed, distributed, and used in approximately equal amounts. The use of MFT may delay the emergence and spread of drug resistance and improve long-run clinical outcomes [29]. To examine these two drug policy interventions, the baseline parametrization of current treatment distribution was modified to include private-market elimination, MFT, or both. We investigated scenarios where immediate or gradual (phased in over ten years) implementation of MFT and private market elimination occurred (S1 Table). A gradual phase-in of a policy is done via ten equal annual reductions in private-market drug use until all private market drugs are eliminated, or annual equal reductions in AL (with a corresponding increase of DHA-PPQ usage) over a ten-year period until AL and DHA-PPQ are used equally in the population (S1 Table).

The permutations of private market elimination, MFT, and implementation timelines resulted in nine drug policy scenarios examined; five additional MFT scenarios with uneven (non-uniform) drug distribution were included (S1 Table). These five remaining drug policy scenarios were designed to evaluate the impact of various MFT mixes of AL and DHA-PPQ, ranging from 100% AL to 50% AL by 10% reductions in AL usage. Since the intent of these scenarios is to evaluate the impact of the MFT drug mixture on the emergence of resistance, rapid private market elimination was used for all of them.

## Results

### Baseline Configuration (de novo emergence)

Under the baseline business-as-usual parameterization, we found that by model year 2038 the national median frequency of 580Y would be 0.142 (IQR 0.135 - 0.152), with a treatment failure rate of 13.11% (IQR 13.03% - 13.18%), although on a local basis (i.e., province or cellular) the values may be higher or lower, the results depending strongly on the stochasticity of initial emergence (Fig. 3). On a provincial basis, Ganzourgou Province had the highest 580Y frequency in 2038 with a median frequency of 0.156 (IQR 0.148 – 0.165) with *Pf*PR_2-10_ ranging from 18.97% to 39.05% (mean of 30.91%) and an under-5 treatment rate of 87%. In general, provinces that have moderate to high *Pf*PR_2-10_ and high ACT usage appear to be at higher risk of accelerated mutation fixation. In order to assess the sensitivity of the model to the mutation rate, the mutation rate was varied to be 5x or 10x faster than the calibrated baseline mutation rate, which resulted in accelerated fixation of drug resistance markers (Fig. 4). Likewise, reducing the mutation rate to be 5x or 10x slower resulted in a lower overall frequency of drug-resistant genotypes, consistent with the reduced rate of mutation (S1 File §8).

**Fig. 3:**
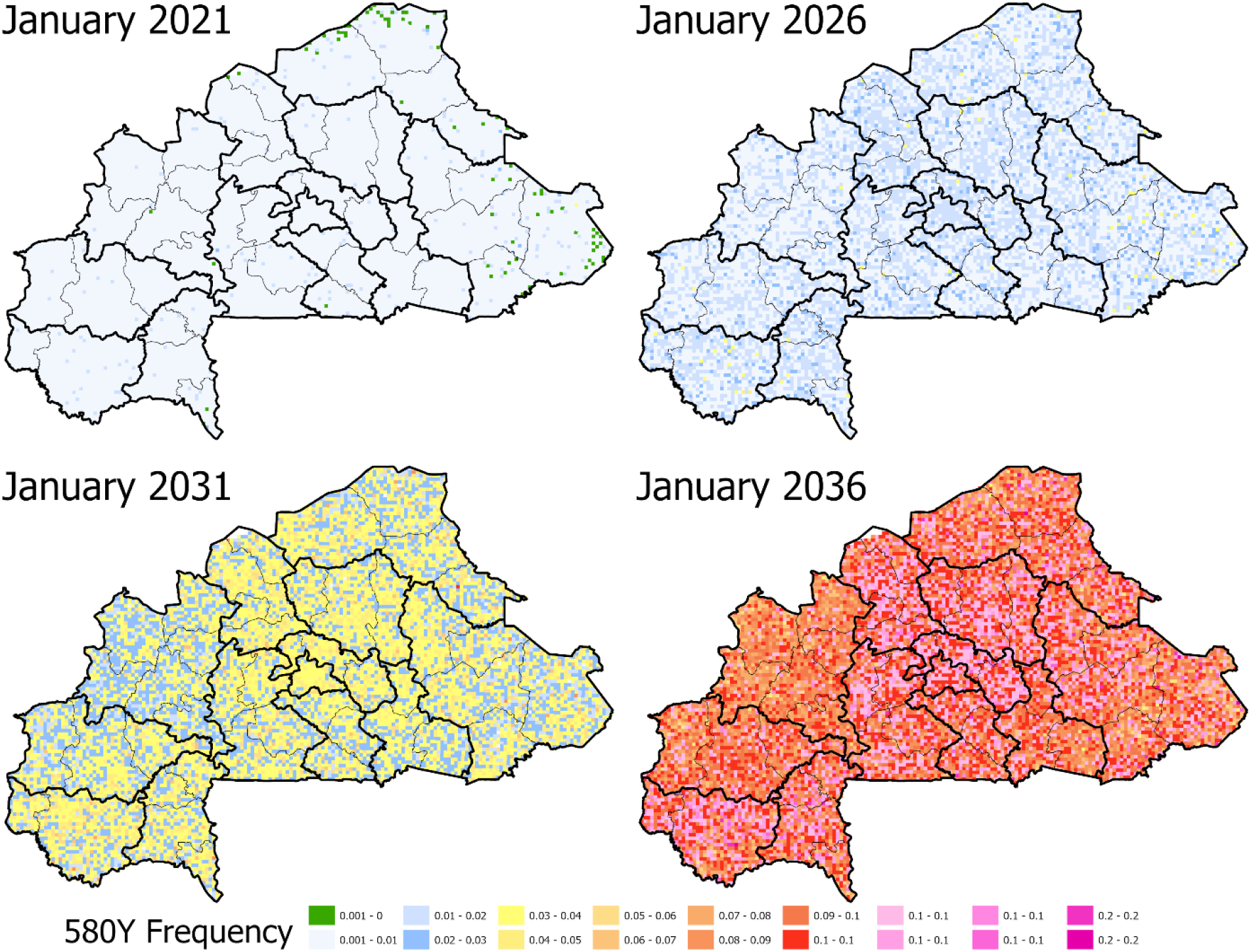
*De novo* emergence of artemisinin resistance under the baseline, status quo scenario. In the baseline scenario, C580 is allowed to mutate to 580Y using a mutation rate that has been set to a previously calibrated pattern of artemisinin resistance in the presence of widespread ACT usage. The appearance of artemisinin resistance in model year 2021 appears to be stochastic with only minimal evidence of elevated frequency of the mutation and some cells with no occurrences. However, by model year 2036 the highest frequencies of artemisinin resistance correspond to areas with elevated to high transmission and high access to treatment. Burkina Faso administrative boundaries from the World Bank Group [19].

**Fig. 4.**
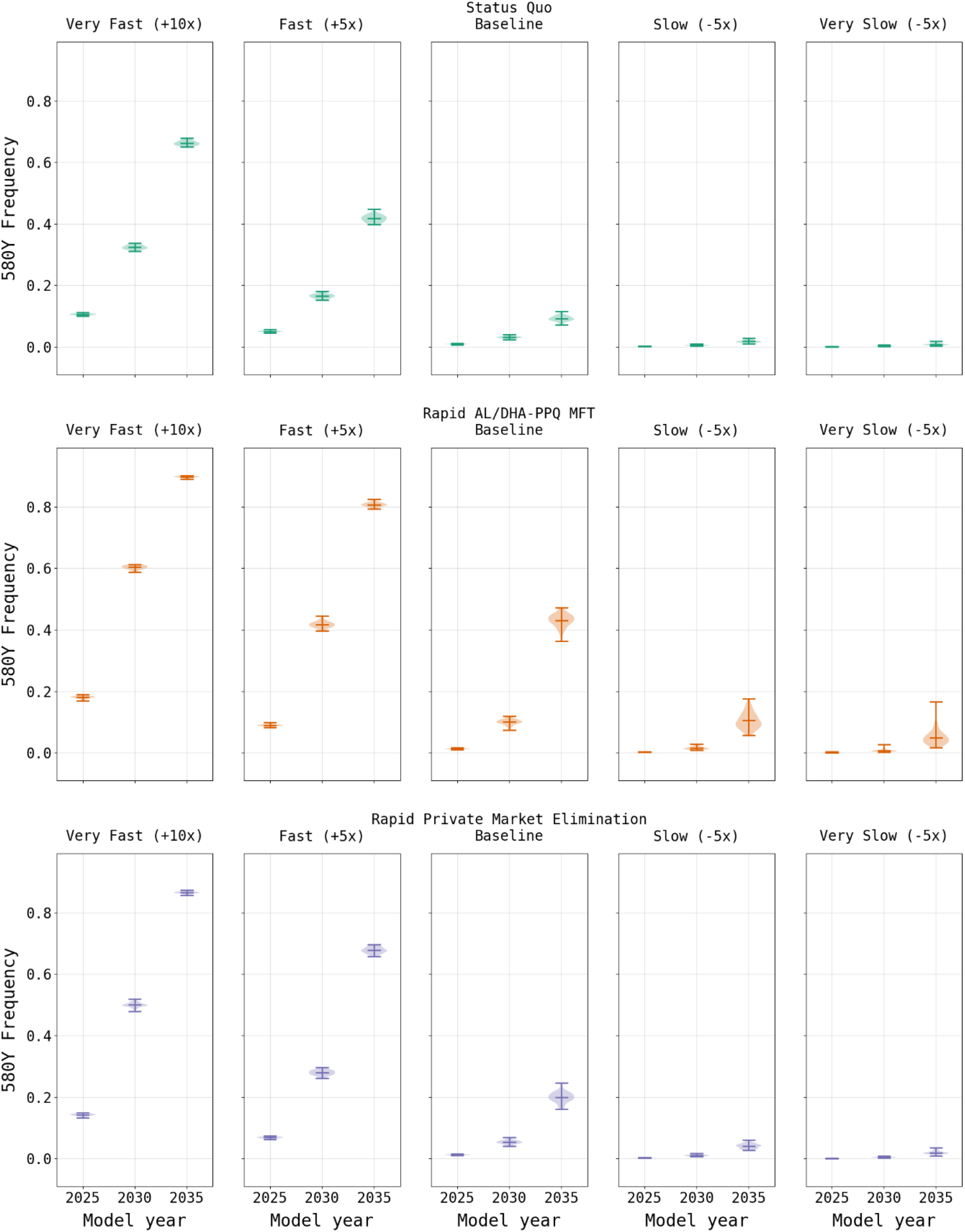
Effects of changes to the mutation rate on 580Y frequency. As expected, the frequency of 580Y is higher when the mutation rate increases to be 5x and 10x faster than the baseline (left two columns) and is lower than the baseline calibrated rate (center column) as the mutation rate decreases to be 5x and 10x slower than the baseline (right two columns).

The spatial distribution of 580Y in the model simulations confirms that (*i*) emergence is stochastic but with relatively low variance, (*ii*) “hot spots” of elevated drug resistance are likely during establishment of drug restraint genotypes, and (*iii*) under high prevalence conditions, high treatment coverage contributes to the acceleration of local selection for the 580Y mutation (or another *kelch13* mutation with similar effect on artemisinin efficacy). Accordingly, the highest frequencies of 580Y under the baseline parametrization occur in the south-western Sudanian climate range (i.e., longer peak transmission season and high treatment coverage), central region (i.e., lower prevalence and high treatment coverage), and northern regions (i.e., high prevalence and high treatment coverage) of Burkina Faso.

### Drug Policy Interventions

As expected, a focus solely upon the elimination of the private market results in an increase in the frequency of 580Y due to the resulting increase in ACT usage (Fig. 5). Under the rapid elimination scenario, the national median frequency of 580Y rises to 0.323 (IQR 0.311 - 0.339) by model year 2038, while under a ten-year elimination strategy the median 580Y frequency in 2038 is 0.266 (IQR 0.254 - 0.279). Elimination of the private market has a favorable outcome on treatment failures due to their reduction in relation to the baseline scenario, with a rapid elimination resulting in a failure rate of 12.90% (IQR 12.75% - 13.09%) in model year 2038 and a ten-year phase out resulting in a slightly lower rate of 11.98% (IQR 11.81% - 12.15%). This reduction in treatment failures in comparison to the baseline configuration, despite the higher 580Y frequency, highlights the value in ensuring that individuals use highly efficacious treatments when presenting with malaria. Note that the model predicts that the intermediate option (i.e., 10-year phase-out) is the best long-term approach for reducing treatment failures, highlighting the combined effects of artemisinin-resistance evolution and high-efficacy treatment access that need to be considered in long-term drug-resistance planning.

**Fig. 5:**
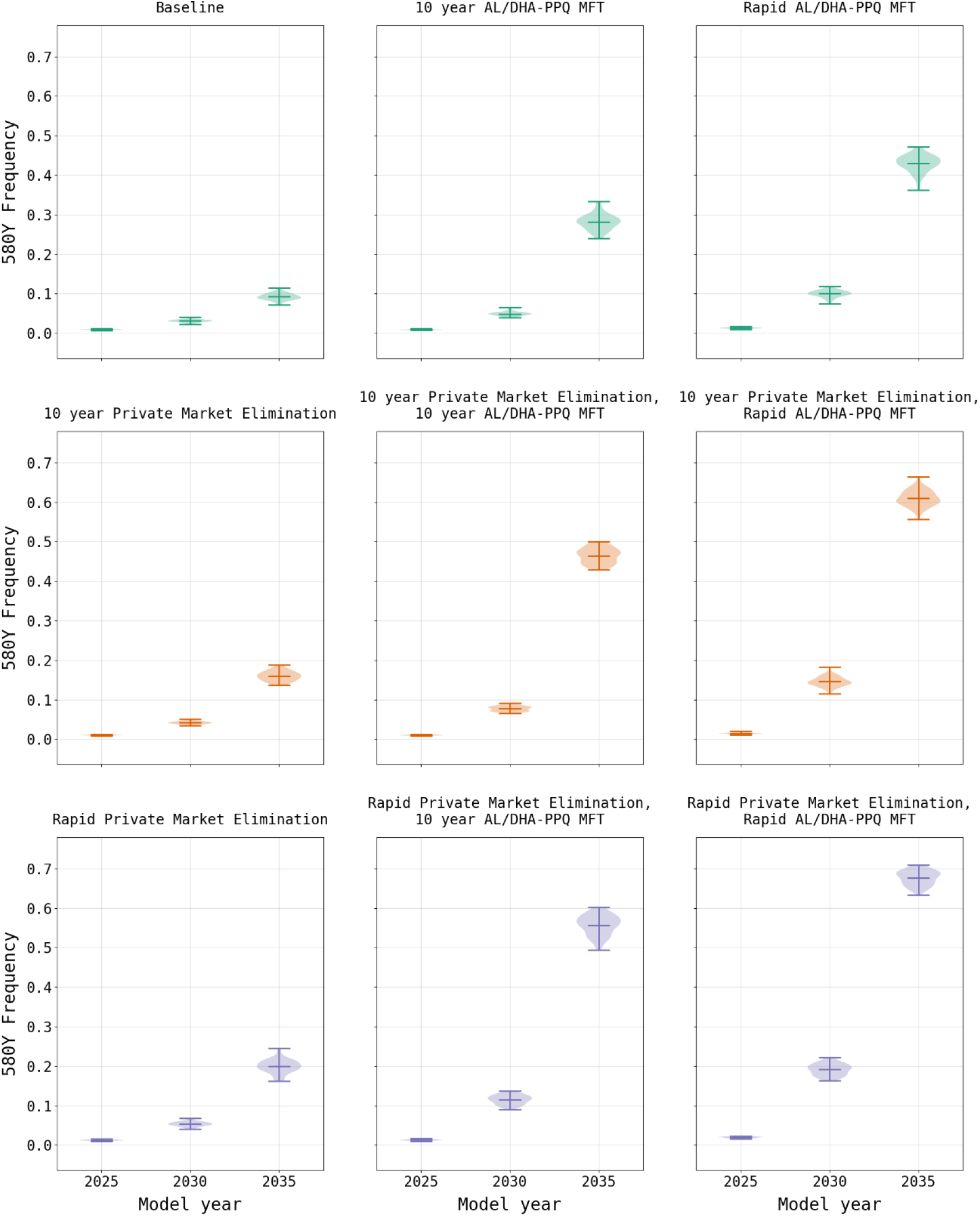
Comparison of 580Y allele frequency (mean and range) under two primary drug policy scenarios. AL/DHA-PPQ MFT refers to a policy of multiple first-line therapies where AL and DHA-PPQ are deployed equally in the population. Evaluations of MFT and private market elimination are shown for rapid (i.e., immediate) implementation and gradual implementation over a 10-year period. Increased use of ACTs due to private market elimination results in a small increase in the frequency of 580Y. Implementation of MFT results in higher long-term 580Y frequencies due to more rapid 580Y evolution enabled by PPQ-resistant parasites.

In contrast to the elimination of the private market, the model suggests that the introduction of MFT with an equal distribution of AL and DHA-PPQ would have a negative effect on treatment failure rates due to a more rapid acceleration in 580Y frequency (Fig. 6), driven primarily by DHA-PPQ. When an MFT strategy with AL and DHA-PPQ is implemented without elimination of the private market, treatment failures increase to a national median of 29.28% (IQR 28.95% - 29.50%) by model year 2038 under immediate implementation conditions, and 26.69% (IQR 26.34% - 26.93%) when MFT is phased in over ten years. While the frequency of 580Y is high under both scenarios, reaching a national median of 0.620 (IQR 0.611 - 0.633) by model year 2038 with rapid adoption and 0.470 (IQR 0.453 - 0.482) with a ten-year introduction, it is insufficient to account for the treatment failure rate alone suggesting multiple factors at play. The rate of treatment failures is accelerated when MFT is coupled with private market elimination. Under a rapid private market elimination scenario, coupled with the immediate adoption of MFT, the resulting national median treatment failure rate in model year 2038 of 34.00% (IQR 33.84% - 34.09%) and 580Y frequency of 0.837 (IQR 0.825 - 0.842) by the end of the simulation. This pattern of increased 580Y frequency is observed across all permutations of private market elimination and MFT adoption rates (Table 1; S2 Table).

**Table 1.** Drug policies examined in this study. Note that all policy interventions result in a higher 580Y frequency compared to the status quo scenario, but this may be offset by reductions in the treatment failure rate for some policies. Note that the MFT scenarios assume rapid private market (PM) elimination and that unless otherwise specified the MFT is 50% AL and 50% DHA-PPQ.

| Drug Policy Studies | 580Y Frequency in 2038 |  | Treatment Failure Rate in 2038 |  |
| --- | --- | --- | --- | --- |
|  | Median | IQR | Median | IQR |
| Status Quo | 0.142 | 0.135 – 0.152 | 13.11% | 13.03% - 13.18% |
| Rapid AL/DHA-PPQ MFT | 0.62 | 0.611 – 0.633 | 29.28% | 28.95% - 29.50% |
| Rapid AL/DHA-PPQ MFT, Rapid PM Elimination | 0.837 | 0.825 – 0.842 | 34.00% | 33.84% - 34.09% |
| Rapid AL/DHA-PPQ MFT, 10 Year PM Elimination | 0.79 | 0.785 – 0.801 | 33.00% | 32.84% - 33.15% |
| 10 Year AL/DHA-PPQ MFT | 0.47 | 0.453 – 0.482 | 26.69% | 26.34% - 26.93% |
| 10 Year AL/DHA-PPQ MFT, Rapid PM Elimination | 0.758 | 0.744 – 0.769 | 32.62% | 32.19% - 32.83% |
| 10 Year AL/DHA-PPQ MFT, 10 Year PM Elimination | 0.687 | 0.671 – 0.697 | 30.96% | 30.67% - 31.16% |
| Rapid PM Elimination | 0.323 | 0.311 – 0.339 | 12.90% | 12.75% - 13.09% |
| 10 Year PM Elimination | 0.266 | 0.254 – 0.279 | 11.98% | 11.81% - 12.15% |
| MFT (60% AL, 40% DHA-PPQ) | 0.761 | 0.752 – 0.775 | 29.48% | 29.35% - 29.75% |
| MFT (70% AL, 30% DHA-PPQ) | 0.65 | 0.639 – 0.664 | 24.66% | 24.42% - 24.90% |
| MFT (80% AL, 20% DHA-PPQ) | 0.507 | 0.489 – 0.524 | 19.55% | 19.24% - 19.81% |
| MFT (90% AL, 10% DHA-PPQ) | 0.353 | 0.335 – 0.367 | 14.07% | 13.81% - 14.26% |
| AL Only, Rapid PM Elimination | 0.342 | 0.321 – 0.35 | 13.23% | 13.05 – 13.79% |

**Fig. 6:**
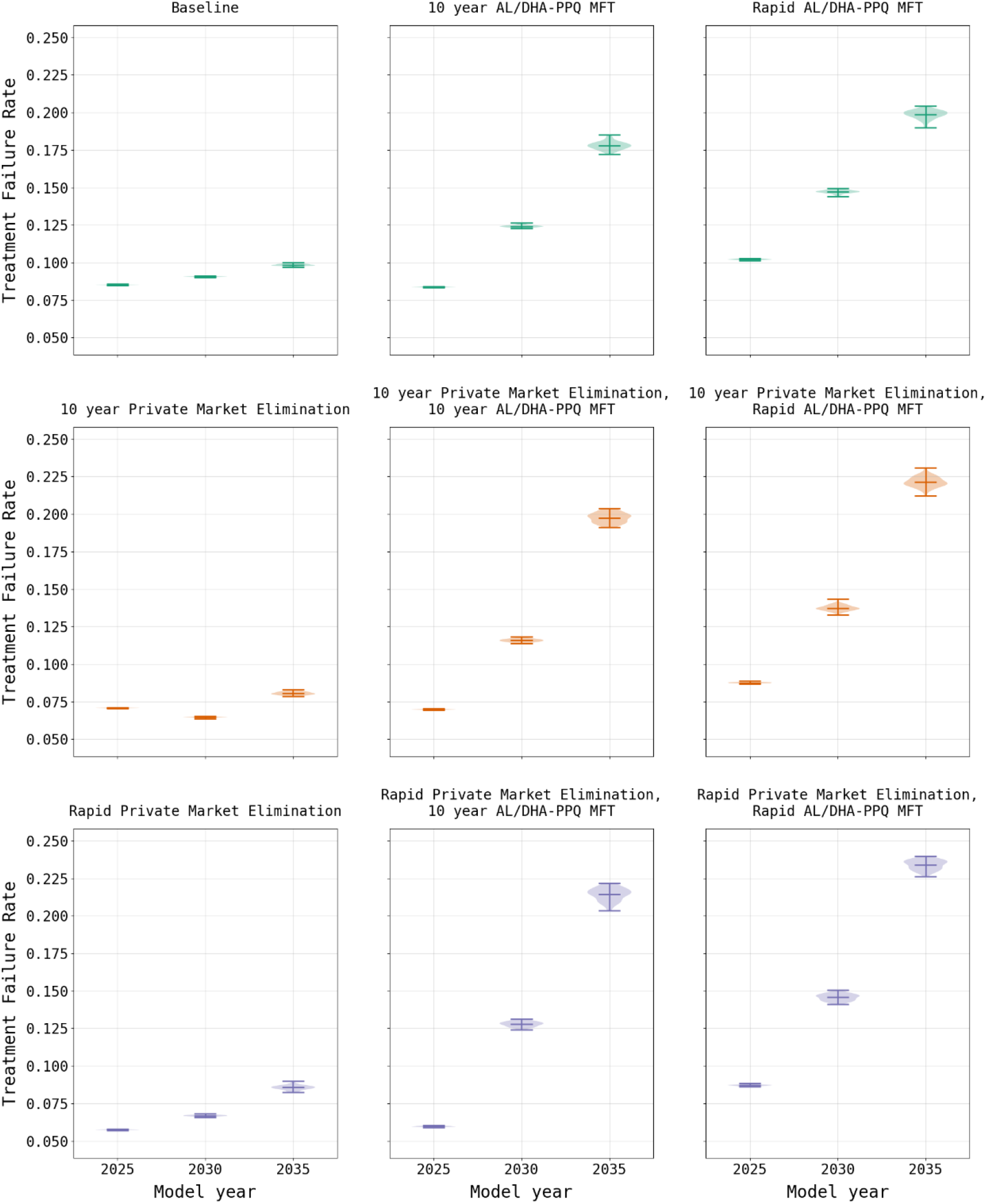
Comparison of treatment failure rates (mean and range) under two policy scenarios. AL/DHA-PPQ MFT refers to a policy of multiple first-line therapies where AL and DHA-PPQ are deployed equally in the population. Evaluations of MFT and private market elimination are shown for rapid (i.e., immediate) implementation and gradual implementation over a 10-year period. Under the baseline scenario the treatment failure rate remains fairly consistent over a 10-year period. With elimination of the private market, the treatment failure rate drops, which is expected given that this approach reduces usage of less efficacious treatments. Introduction of MFT together with elimination of the private market also reduces usage of less efficacious treatments; however, this is rapidly offset by the increased treatment failure rate due to the increasing frequency of *plasmepsin-2,3* double-copy genotypes.

This adverse MFT outcome is attributable to the increased usage of DHA-PPQ as the co-equal first-line therapy. Typically, MFT policies result in better long-term drug-resistance outcomes due to the heterogeneous drug environments they create [29,30]. However, these analyses assume that all therapies distributed in an MFT policy are equally efficacious and have identical drug-resistance properties. This is not the case when comparing DHA-PPQ and AL, as piperaquine (PPQ) resistance leads to higher levels of treatment failure when compared to lumefantrine resistance [24]. In contrast to the baseline scenario with limited DHA-PPQ usage (5.1%), or the nominal increase resulting from private market elimination (6.2%), introduction of MFT results in DHA-PPQ usage in 46.1% of treatments when coupled with private market elimination (S1 Table). The drug policy combination of rapid private market elimination along with rapid MFT introduction results in an accelerated frequency increase of the *plasmepsin-2,3* double-copy genotypes conferring PPQ resistance [31]. Effectively, following acquisition of PPQ resistance, the continued selective pressure of DHA produces an environment that is more favorable to the evolution of artemisinin resistance due to the lack of partner-drug killing of potentially emergent artemisinin-resistant genotypes. This is consistent with recent findings that partner-drug resistance facilitates the development of primary drug (i.e., artemisinin) resistance [25]. The increased frequency of *plasmepsin-2,3* double-copy genotypes is observable shortly after implementation of the policy in model year 2021 (Fig. 7) and is present in all scenarios that implement MFT (Fig. 8) as well as under sensitivity analysis when a reduced mutation rate is used (S1 File §8).

**Fig. 7:**
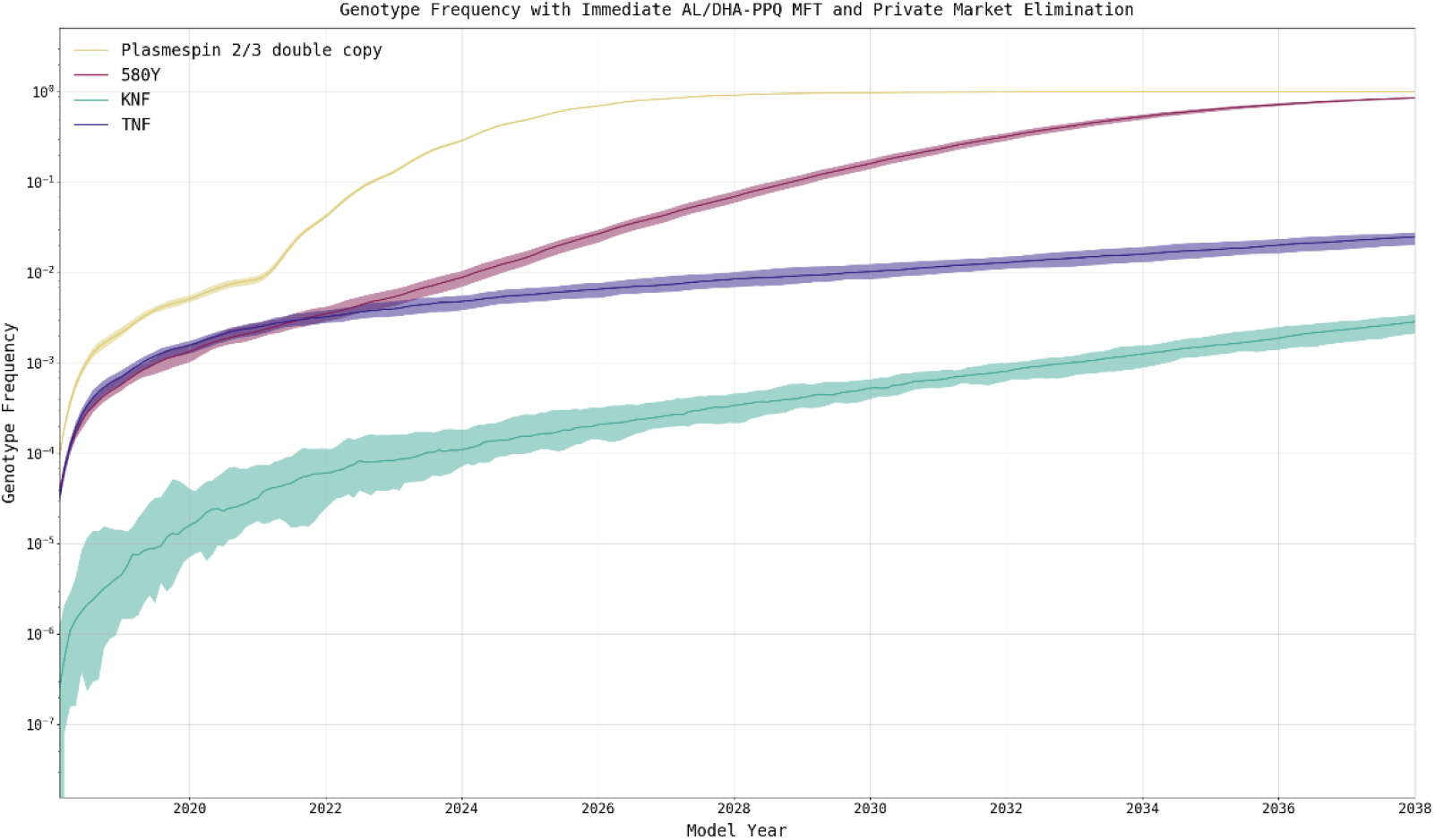
Increase of *plasmepsin-2,3* double-copy genotypes under immediate private-market elimination and immediate MFT (median and 95% range). The rapid elimination of the private market coupled with introduction of MFT (in model year 2021) serves as an exemplar of the rapid increase in double-copy *plasmepsin-2,3* genotypes after DHA-PPQ is introduced at high levels in 2021. Double-copy *plasmepsin-2,3* genotypes (yellow) are strongly selected for after DHA-PPQ is introduced, which contrasts with the slower pace of 580Y evolution (magenta). This rapid increase in the frequency of the double-copy *plasmepsin-2,3* genotypes result from the low efficacy of DHA-PPQ on these genotypes (77% efficacy on C580 and 42% on 580Y). KNF genotypes (green) show the most resistance to lumefantrine, but the efficacy of AL on KNF genotypes is 89% (C580) or 72% (580Y) making the selection pressure exerted by AL weaker than the selection pressure exerted by DHA-PPQ.

**Fig. 8:**
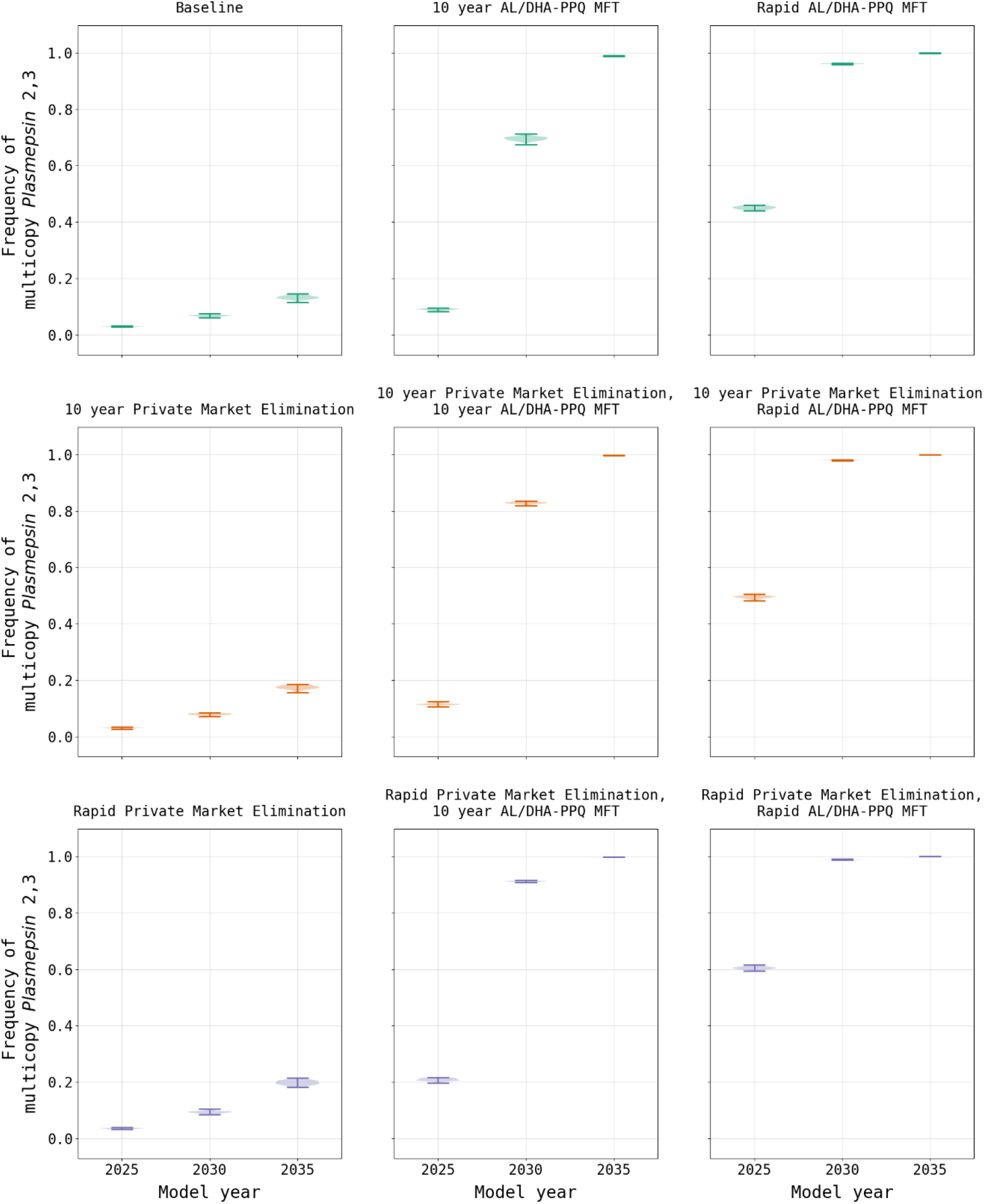
Comparison of the modeled distribution of *plasmepsin-2,3* double-copy frequency under two drug policies. AL/DHA-PPQ MFT refers to a policy of multiple first-line therapies where AL and DP are deployed equally in the population. Evaluations of MFT and private market elimination are shown for rapid (i.e., immediate) implementation and gradual implementation over a 10-year period. Under policies of private market elimination, there is little effect on DHA-PPQ usage and thus low pressure on *plasmepsin-2,3* evolution. When MFT is introduced, the usage of DHA-PPQ increases substantially, resulting in rapid selection of double-copy *plasmepsin-2,3* genotypes.

To further investigate the parameters under which an MFT policy incorporating DHA-PPQ may be implemented in Burkina Faso, additional scenarios were considered in the absence of a private market: the use of AL as the sole first-line therapy, along with MFT combinations of AL and DHA-PPQ including a range from 60% to 90% AL use (S1 Table). When AL is used as a sole first-line therapy for uncomplicated malaria, 580Y frequency reaches 0.342 (IQR 0.321 - 0.350) by model year 2038 with a treatment failure rate comparable to that of the status quo scenario of 13.23% (IQR 13.05% - 13.37%). When MFT is introduced using DHA-PPQ (10% of treatments) and AL (90%) the results by model year 2038 show a slight increase in the 580Y frequency (0.353; IQR 0.335 - 0.367; p < 0.0001; Wilcoxon Rank-Sum) and treatment failure rate (14.07%, IQR 13.81% - 14.26%, p < 0.0001) when compared to the AL as the sole first-line therapy. When the usage of DHA-PPQ is increased to 20% or higher, there are further increases in 580Y frequencies along with treatment failure rates by model year 2038 (Table 1, Fig. 9).

**Fig. 9:**
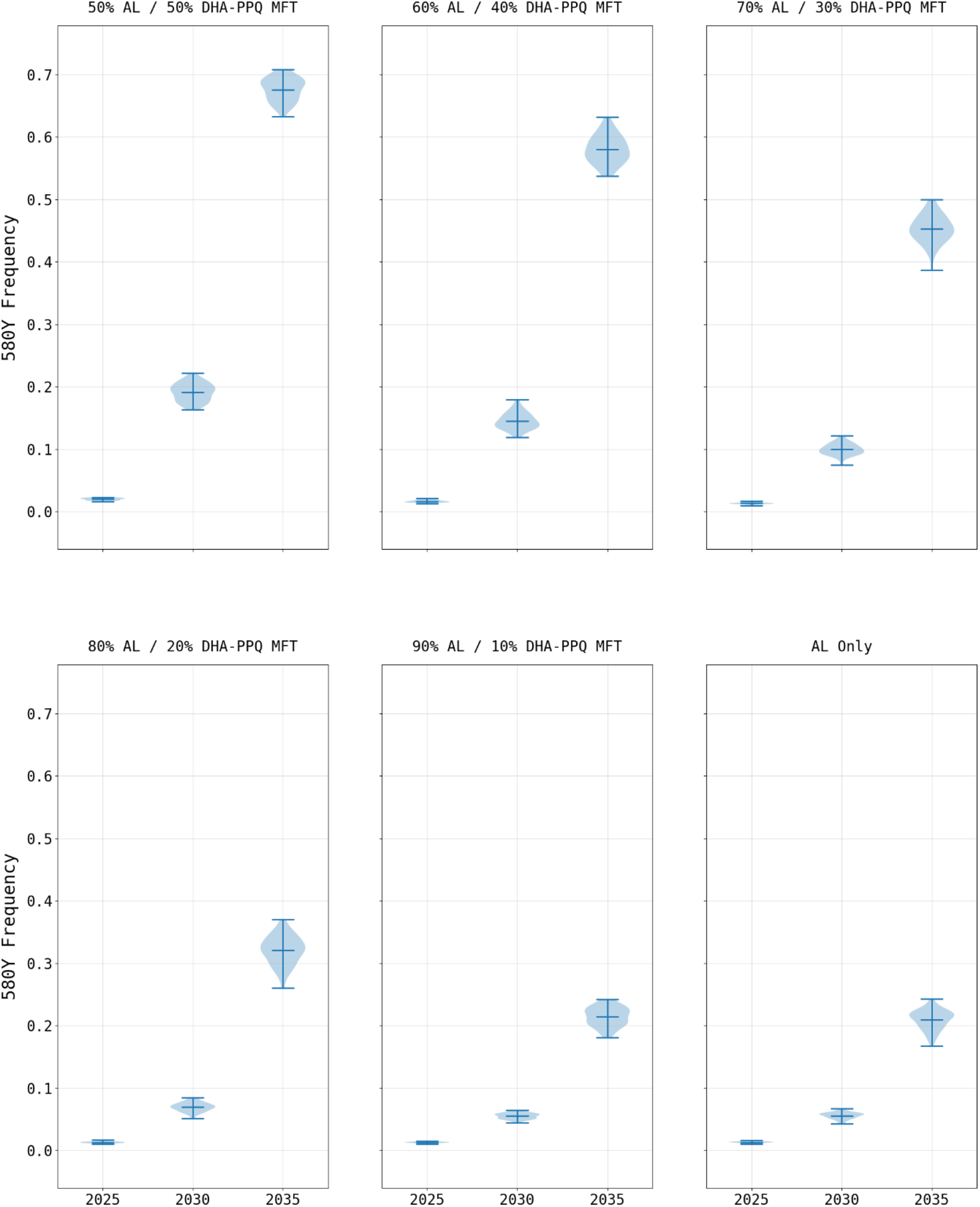
580Y frequency under various MFT configurations (mean and range). The impact of changing the role of DHA-PPQ in an MFT policy. Lower DHA-PPQ use results in weaker selection pressure on 580Y alleles. The loss of PPQ efficacy due to *plasmepsin-2,3* double-copy emergence influences the rate at which the 580Y frequency increases.

### Seasonal changes in alleles frequencies

As in some observed field studies [32,33], drug-resistance allele frequencies in our model fluctuated with the periodicity of the transmission season. This effect is most pronounced in regions of Burkina Faso where there is highly seasonal transmission (e.g., northern Sahelian climate zone). In order to examine this effect, additional simulations where conducted that reduced the geographic scope of the model to a single 25 sq.km cell, allowing for more model detail to be captured (S1 File §9). Our simulations show an increase in the ratio of symptomatic/asymptomatic individuals over the course of the rainy season, resulting in increased evolutionary pressure favoring drug resistance due to a higher proportion of *P. falciparum* infections being treated. The reason for the increase in the symptomatic/asymptomatic ratio during peak transmission likely results from several factors, namely, (*i*) a lower population-average level of immunity at the beginning of the high transmission season, (*ii*) possibility for multiple bites and repeat symptoms for individuals with low immunity, and (*iii*) correlation between low biting attractiveness and low immunity which will skew the ratio of bites on immunes/non-immunes from low to high transmission periods. Although the seasonal trend in drug-resistance frequency was up during the rainy season and down or flat during the dry season, the overall long-term trend remained upward with increasing drug resistance.

## Discussion

While the evolution of drug resistance by the *P. falciparum* parasite is a pressing concern, in HBHI countries drug resistance evolution is secondary to the need to increase access to and usage of ACTs. Once access to ACTs is near universal, more attention should be turned towards the future impacts of drug resistance. As noted by Valle et al. [34], the spatial distribution of drug access and disease burden cannot be presumed to be uniform, and this heterogeneity will have onward effects on which areas need more focus on basic public health access versus drug-resistance management. As policy makers need to work within their local constraints when designing national drug policies, the overall national-scale timeline of the emergence and spread of drug resistance should still be the main focal point in the preparation and design of control strategies for drug resistance.

Presently there are early signals of ACT treatment failures starting to appear in Burkina Faso [13,14], although a genetic mechanism has not been identified, nor is it clear how widespread these signals are. If these signals are confirmed to have a genetic basis, then our study suggests that widespread resistance may manifest within a decade if there are no changes to drug policies. While the present drug mixture in Burkina Faso results in a low amount of evolutionary pressure on the parasite (specifically, pressure on artemisinin resistance via the 580Y allele), the rates of ACT usage are likely to increase in the future [23], indicating that evolutionary pressure will strengthen as access to ACTs increases and other interventions are implemented. Given the negative impact the private market has upon the overall treatment failure rate, this study suggests policy makers should explore ways to reduce or eliminate the use of private market drugs. Doing so would ensure broader use of highly efficacious ACTs and have a minimal impact upon the projected rise of drug resistance given the current drug mixture in Burkina Faso.

This study highlights the need for ACT partner-drug resistance awareness, particularly when partner-drug resistance has already been identified [35]. In the long term, drug resistance is expected to evolve in all scenarios where population-level drug-use is sufficiently high, but resistance does not imply complete loss of efficacy. For example, in the case of lumefantrine, the efficacy of AL remains high at 89% on certain lumefantrine-resistant genotypes [24]; however, DHA-PPQ efficacy drops to 77% when PPQ resistant parasites are present [24]. Ultimately, the partner drug moderates the pace at which artemisinin resistance can emerge. Rapid acquisition of resistance to the partner drug results in an acceleration of artemisinin resistance evolution due to the usage of ACTs as a ‘de facto monotherapy’ because of low efficacy of the partner drug.

Despite the increase in drug resistance and treatment failures under a balanced MFT approach, elimination of the private market (with or without MFT) may offer policy makers some flexibility in managing future drug-resistance concerns. If MFT is not implemented, then this study suggests that the overall rate of treatment failures will go down as the public switches to more efficacious treatments. The selective pressure on the parasite will increase with more public-sector drug use, and this is assumed in the model regardless of intervention due to yearly efforts to increase access to ACTs. Furthermore, this study suggests that the timeframe for gaining benefits from private market elimination may be quite broad. While complete private-market elimination is unlikely outside of a model, phasing out the private market over ten years may be achievable.

Any modeling exercise is dependent upon assumptions and simplifications that may impact the results. The primary limitation of this model is that it does not account the 2018 introduction of a broad SMC program intended to reach all children under-5 in Burkina Faso [12]. Given the recency of this broad approach to the program, limited data are available to calibrate model parameters accurately. The exclusion of SMC in our analysis likely results in a simulated prevalence that is higher than what is likely to be observed in the field. However, this needs to be moderated against the unknown impacts of the ongoing COVID-19 pandemic, which may impact malaria control programs and possibly lead to higher *Pf*PR_2-10_ than what was used for model calibration, although the real impacts may not be quantifiable for several years [36,37].

While the mutation rate used by the simulation is based upon an accepted alignment based on the observed pattern of *kelch13* evolution in Southeast Asia over the past two decades [25], the true mutation rate may be faster or slower. This is a common limitation of mathematical models of evolution since values for mutation parameters are difficult to estimate during short periods of observation [38,39]. As such, the mutation rates result in simulation outputs wherein the relative results can be compared (i.e., private market elimination versus status quo) but the absolute values are susceptible to error if the mutation rate assumption is incorrect. Another limitation is the simplified nature of the pharmacokinetic model that is being used. While an individual multi-compartment model for each compounded incorporated in the simulation would be the best approach [40], the complexity of implementation was balanced against the objectives of this simulation, namely, to ensure that the parasite killing rate and evolutionary pressure resulting from treatment was consistent with the 24-hour timestep used by the simulation. It is not known how model misspecification of PK/PD dynamics on a minute/hour timescale affects long-term evolutionary outcomes years or decades into the future.

It has been well established that asymptomatic individuals with *P. falciparum* play an important role in the propagation of the parasite between rainy seasons [41]; however, the impact that this has upon the evolution of drug resistance has not been well established. The seasonal periodicity of resistance markers that appeared in this simulation, in conjunction with previous results concerning the role that seasonality and asymptomatic carriers may play in the selection of drug-sensitive or drug-resistant parasites [32,41], suggests that more work is needed to understand how seasonal transmission affects selection. Improved simulation approaches will be necessary to assist policymakers in addressing the critical question of when ACTs will need to be replaced? While ACTs remain highly effective in Africa, this study, along with recent reports on treatment failure [7,13,14] suggest that we may be starting to enter a period of emerging artemisinin resistance and an end to the current “calm before the storm” suggested by Conrad and Rosenthal [42]. This transition is inevitable. ACT access is likely to increase in the coming years, reducing case numbers but increasing the selection pressure of artemisinin-resistant genotypes. Preparation, prevention, and preemption during this high-risk period for drug-resistance emergence will be key to ensuring that first-line treatment against *P. falciparum* continue working at high efficacy through the 2020s and 2030s.

## Data Availability

Source code and intermediate data files are available on GitHub.

https://github.com/bonilab/malariaibm-spatial-BurkinaFaso-2021

## Data availability

Spatial raster files used in the simulation and long with intermediary files used for analysis and plot generation can be found at https://github.com/bonilab/malariaibm-spatial-BurkinaFaso-2021/tree/main/Data

## Code availability

The source code for the base mathematical model and analysis specific to this manuscript https://github.com/bonilab/malariaibm-spatial-BurkinaFaso-2021

## Acknowledgements

Simulations described in this study were performed on the Pennsylvania State University’s Institute for Computational and Data Sciences’ Roar supercomputer.

## Author Contributions

RJZ developed the model’s spatial framework, fit the model to movement and epidemiological data, conducted the simulation studies, prepared figures, maps, and drafted the manuscript. RJZ and TDN integrated the spatial framework into the previous model version and carried out epidemiological validation analyses. KTT performed a code review of the simulation and assisted in debugging. AFS, JG, AW, J-BO guided the calibration of the model’s epidemiological and movement behaviors. TN-AT parameterized the model’s partial drug-resistance phenotypes and evaluated these phenotypes’ trajectories in the model simulations. PD developed visualizations for model outcomes. DDHG developed a pilot model for testing. RJZ and MFB designed the simulation study and edited the manuscript. All authors read and approved the final version of the manuscript.

## Supporting Information

**S1 File. Supplemental Materials**. This PDF file contains additional information concerning the model parameterization, calibration, validation, and sensitivity analysis.

**S1 Table. Studies performed and treatment configuration**. This Excel file contains a complete list of the studies performed along with the treatments used.

**S2 Table. Complete 580Y frequency and treatment failure rates**. This Excel file contains the complete list of studies performed along with the 580Y frequency and treatment failure rates.

