## Supplementary material for "Long-term effects of increased adoption of artemisinin combination therapies in Burkina Faso": S1 File

---

### 1 Model Spatial Structure

While the simulation supports arbitrary cell sizes, a 5km-by-5km (25 sq.km) cell size was selected since it corresponds to the cell size of the *Pf*PR$_{2\text{-}10}$ reference data from the Malaria Atlas Project (Weiss et al. 2019). This results in the simulation having 10,936 cells of data (or simulation cells) with a total of 126 rows by 173 columns (10,862 cells are outside of the national boundaries). During model initialization spatial data is loaded in the form of the national provinces, eco-climatic zones (see Section 2 for more details), population (Section 3), access to treatments (Section 6), travel to the nearest city (Section 4), and the beta (i.e., transmission parameter) for each cell. During model execution results are tracked at the cell level and aggregated up to a provincial, regional, or a national level as needed.

**Figure S1.** Projected annual mean PfPR2-10 by Malaria Atlas Project for 2017 (Weiss et al. 2019), note the cellular resolution of 5km-by-5km. Map prepared by the authors and originally appeared in Zupko et al. (2021), Africa boundaries from OpenStreetMap (2017), Burkina Faso administrative boundaries from World Bank Group (2018).

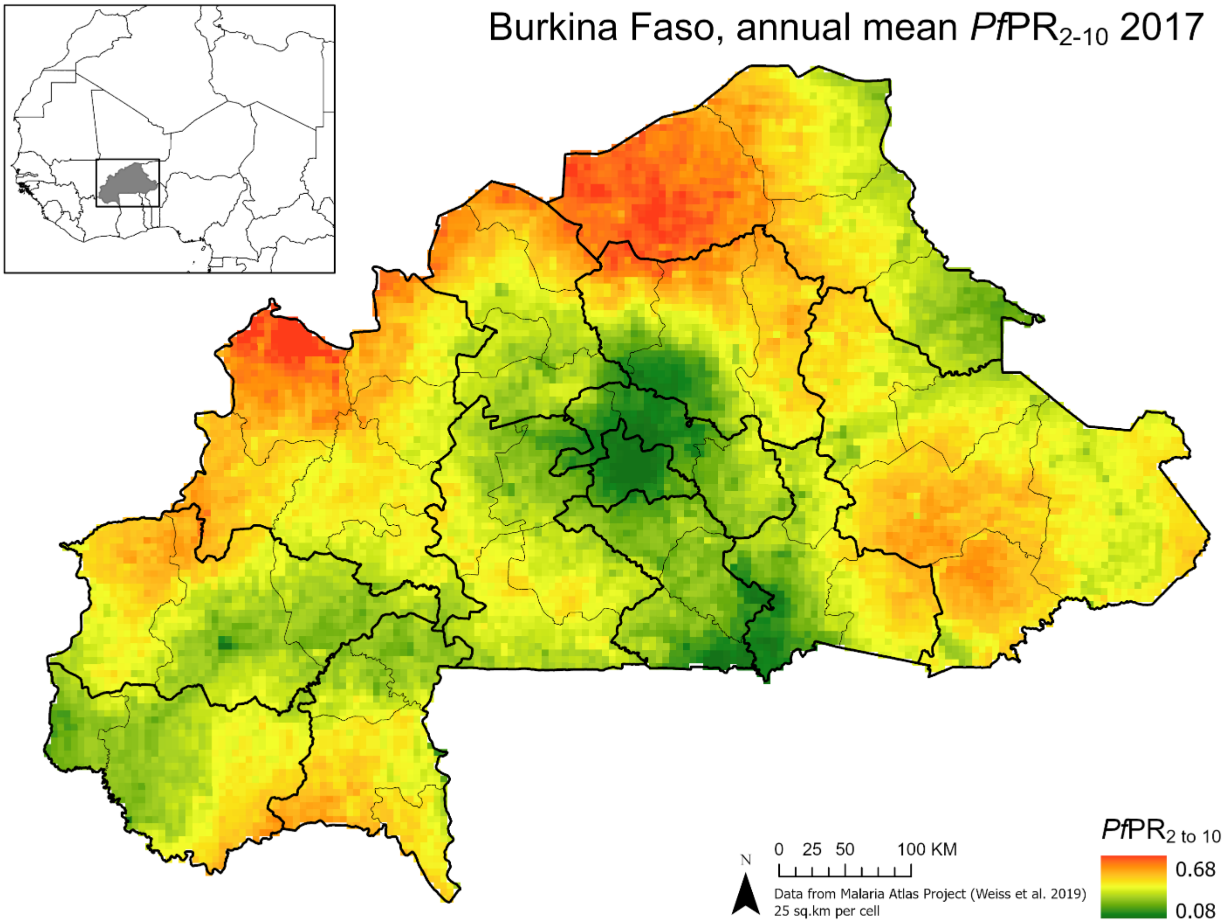

### 2 Seasonality

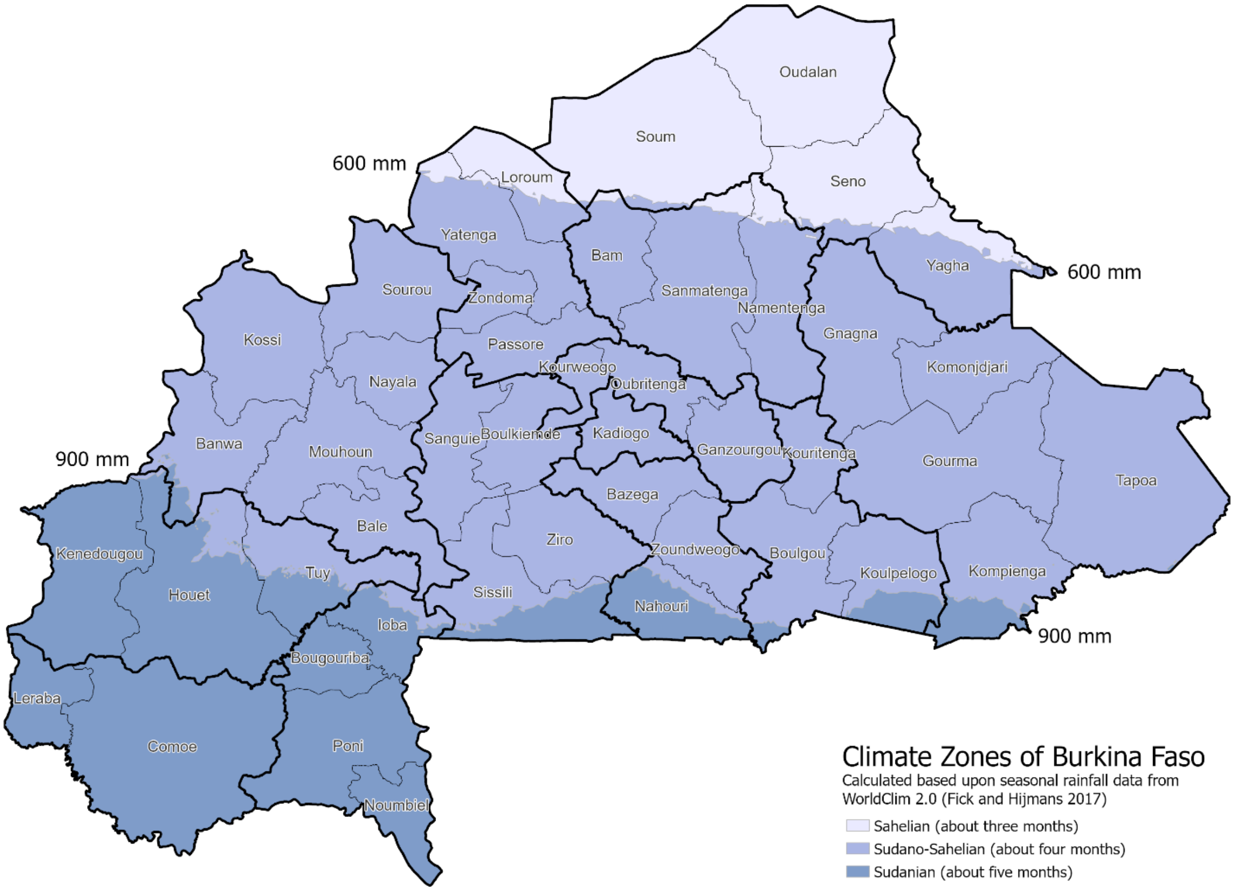

The seasonality of malaria in Burkina Faso approximately corresponds to the eco-climatic zones of the Sudanian, Sudano-Sahelian, and Sahelian regions, which are defined by the rainfall boundaries of 900 mm, and 600 mm per year (Figure S2; Kagone 2006; Fischer et al. 2011). These regions in turn correspond to three (Sahelian), four (Sudano-Sahelian), and five months of transmission (Sudanian). In order to define these regions within the model, monthly WorldClim 2.0 precipitation data (Fick & Hijmans 2017) was aggregated to an annual basis and used to define the geographic boundaries. Following delimitation of the boundaries, parameters were fit for the following equation:

$$multiplier = base + \left(a * \sin^{+}\left(\frac{b * \pi(t - \varphi)}{365}\right)\right) \quad \text{Eq. 1}$$

Where *base* defines the lower bounds for transmission, *a* defines the additive upper bounds, *b* determines the duration of the rainy season, and $\varphi$ determines the peak of the rainy season. The resulting *multiplier* can then be applied to the model's transmission parameter ($\beta$) to produce seasonal variation in transmission intensity. The parameters resulting from the fit (Table S1), produce an approximation of the seasonality patterns for the Sudano-Sahelian, and Sahelian regions (Figure S3).

**Table S1.** Calibration settings for the seasonal prevalence of malaria.

| | **Base** | *a* | *b* | $\varphi$ |
|---|---|---|---|---|
| Sahelian | 0.4 | 0.6 | 2.5 | 146 |
| Sudano-Sahelian | 0.4 | 0.6 | 2.3 | 155 |
| Sahelian | 0.4 | 0.6 | 1.9 | 155 |

**Figure S3.** Modeled seasonality patterns for Burkina Faso. Since the objective of these is to replicate an annual increase and decrease in the transmission of the parasite, the intent is not a precise model of the seasonal curves, but rather a reasonable approximation with similar peaks.

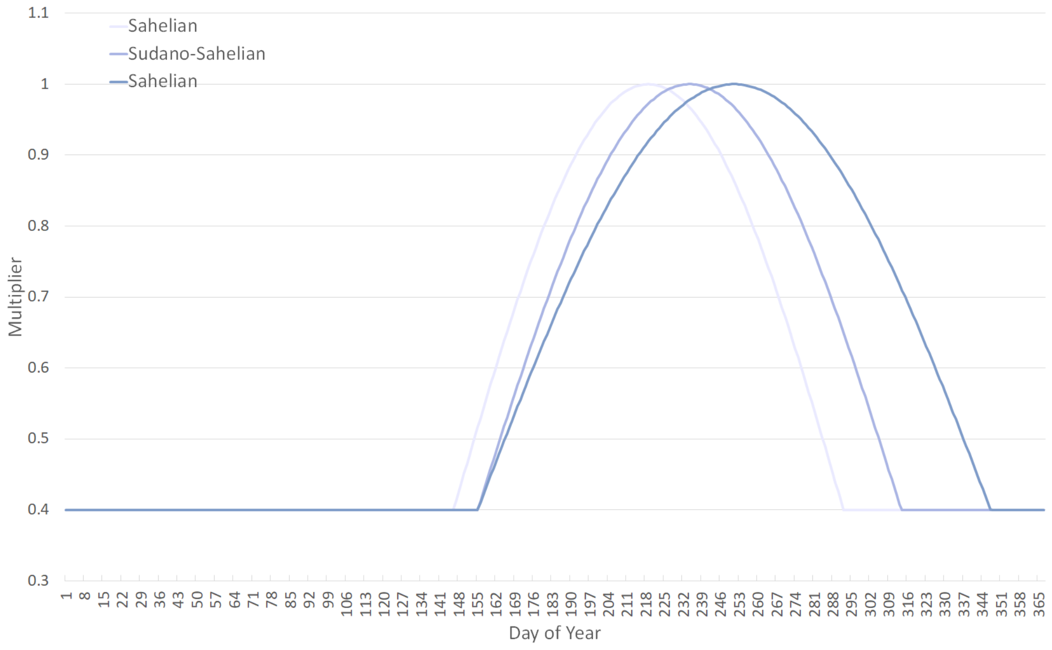

### 3 Demographics and Mortality

**Table S2.** Population distribution of Burkina Faso, using the 2017 estimated population.

| Age Band | Population | Percentage | Age Band | Population | Percentage |
|---|---|---|---|---|---|
| < 5 | 3,525,706 | 18.0% | 45 to 49 | 607,436 | 3.1% |
| 5 to 9 | 3,087,852 | 15.7% | 50 to 54 | 491,293 | 2.5% |
| 10 to 14 | 2,632,326 | 13.4% | 55 to 59 | 386,018 | 2.0% |
| 15 to 19 | 2,189,799 | 11.2% | 60 to 64 | 292,493 | 1.5% |
| 20 to 24 | 1,717,098 | 8.7% | 65 to 69 | 221,482 | 1.1% |
| 25 to 29 | 1,354,144 | 6.9% | 70 to 74 | 160,000 | 0.8% |
| 30 to 34 | 1,101,379 | 5.6% | 75 to 79 | 96,701 | 0.5% |
| 35 to 39 | 940,308 | 4.8% | 80+ | 89,614 | 0.5% |
| 40 to 44 | 738,498 | 3.8% | | | |
| **Total** | | | | **19,632,147** | **100.0%** |

The estimated population of Burkina Faso was 19,632,147 in 2017 (Table S2) and is projected to double in 22 years (INSD et al. 2018, p. 2; INSD 2019). There is some disagreement as to the estimated crude birth rate with figures of 35.1 for 2017 (INSD et al., 2018), 37.5 for 2019 (UN 2019b, p. 334), and 46 in 2006 (INSD 2018, p. 10) appearing in the literature.[1] The model uses a crude birth rate of 37.5 since it gives a reasonable approximation of the 22 year doubling time at 23.76 years assuming a crude death rate of 7.9 (UN 2019a, p. 15).

Since malaria is a leading cause of death for children under-five (Table S3), it is necessary to adjust the mortality rate for the model to exclude deaths due to malaria (Table S4) (UN 2019a). This is done by reducing the all-cause mortality rate for individuals in the simulation by the mortality rate attributable to malaria. Since the model results in mortality due to malaria during the simulation, the all-cause mortality rate achieved *in silico* approximates the recorded all-cause mortality rate.

[1] The crude birth rate of 35.1 for 2017 is provided in USAID Demographic and Health Survey's data, which cites INSD et al. (2018) as the source.

**Table S3.** All-cause, malaria, and non-malaria morality rates, WHO estimated deaths (thousands) (UN 2019a).

| | | All | 0 to 4 | 5 to 14 | 15 to 29 | 30 to 49 | 50 to 59 | 60 to 69 | 70+ |
|---|---|---|---|---|---|---|---|---|---|
| **2016** | All Causes | 159.4 | 59.5 | 13.8 | 16.4 | 19.3 | 10.4 | 13.8 | 26.3 |
| | Malaria | 21.3 | 14.6 | 3.3 | 1.0 | 0.9 | 0.4 | 0.5 | 0.7 |
| | Non-Malaria | 138.1 | 44.9 | 10.5 | 15.4 | 18.4 | 10.0 | 13.3 | 25.6 |
| **2015** | All Causes | 158.9 | 60.7 | 13.9 | 16.3 | 19.3 | 10.2 | 13.5 | 25.1 |
| | Malaria | 22.1 | 15.6 | 3.2 | 1.0 | 0.8 | 0.3 | 0.5 | 0.7 |
| | Non-Malaria | 136.8 | 45.1 | 10.7 | 15.3 | 18.5 | 9.9 | 13.0 | 24.4 |
| **2010** | All Causes | 167.1 | 75.5 | 13.9 | 15.2 | 18.4 | 8.7 | 12.6 | 22.9 |
| | Malaria | 34.3 | 28.7 | 2.9 | 0.8 | 0.7 | 0.3 | 0.4 | 0.5 |
| | Non-Malaria | 132.8 | 46.8 | 11.0 | 14.4 | 17.7 | 8.4 | 12.2 | 22.4 |
| **2000** | All Causes | 187.2 | 99.5 | 13.5 | 15.0 | 18.6 | 7.5 | 11.5 | 21.5 |
| | Malaria | 38.6 | 34.4 | 2.4 | 0.5 | 0.4 | 0.2 | 0.3 | 0.4 |
| | Non-Malaria | 148.6 | 65.1 | 11.1 | 14.5 | 18.2 | 7.3 | 11.2 | 21.1 |

National level statistics for Burkina Faso are used to initialize the simulation upon startup. Individuals are spatially distributed according to the 2019 population map, presuming the age distribution is consistent throughout the county (WorldPop 2018). During initialization, the model generates individuals that are proportionally consistent with the age bands provided for each cell, presuming that one-year subdivisions are evenly divided. In order to account for the 2006 start-date, the population in each cell was multiped by 0.714. This adjustment allows the model to match the expected population by the end of the burn-in period.

**Table S4.** All-cause mortality versus the non-malaria mortality rates used in the model.

| Age | Range | All-cause Mortality (Data) | Non-Malaria Mortality (Model) |
|---|---|---|---|
| 1 | 0 to 4 | 0.05100 | 0.03820 |
| 2 | | 0.04031 | 0.03019 |
| 3 | | 0.02706 | 0.02027 |
| 4 | | 0.02036 | 0.01525 |
| 5 | 5 to 14 | 0.01632 | 0.01248 |
| 6 | | 0.00469 | 0.00359 |
| 7 | | 0.00472 | 0.00361 |
| 8 | | 0.00477 | 0.00365 |
| 9 | | 0.00495 | 0.00379 |
| 10 | | 0.00495 | 0.00379 |
| 11 | | 0.00505 | 0.00386 |
| 15 | 15 to 29 | 0.00628 | 0.00590 |
| 20 | | 0.00707 | 0.00664 |
| 60 | 60 to 69 | 0.01807 | 0.01727 |
| 100 | 70+ | 0.37676 | 0.36621 |

### 4 Individual Movement

The movement of human populations plays a role in the spread of malaria. To account for this, a movement model was developed and validated for our simulation (Zupko et al. 2021), describing movement from pixel to pixel in the simulations' spatial structure that is summarized here as follows. Briefly, summarizing the calibration procedure from Zupko et al (2021), we start with a modified gravity model suggested by Marshall et al (2018):

$$Pr(j|i) \propto Pop_j^{\tau} k(d_{i,j}) \quad \text{Eq. 2}$$

$$k(d_{i,j}) = \left(1 + \frac{d_{i,j}}{\rho}\right)^{-\alpha} \quad \text{Eq. 3}$$

where *i* corresponds to the source locations and *j* to the destination. Probability of travel from origin *i* to destination *j* is proportional to the product of (1) the population at *j* raised to *τ* power and (2) the distance kernel, which takes the form of a power law as shown in Equation 3. The kernel includes the scale parameter $\rho$ and power-law parameter $\alpha$ to allow for a fit to the raw travel data. A friction surface based upon the travel time to the nearest city (Weiss et al. 2019) is then incorporated by adjusting the probability *Pr(j|i)*:

$$Pr' = \frac{Pr}{(1 + t_i + t_j)} \qquad \text{Eq. 4}$$

Where *t* represents the travel time from site $i$ or $j$ to the nearest city. While this adjustment still allows for travel to and from difficult to access parts of the map, the likelihood of travel to low-access (high friction) region is reduced. Finally, a penalty was applied to the capital province Kadiogo for within-province travel, where individuals have their intra-province travel probabilities divided by a factor of 12 to reduce intra-province travel in Ouagadougou and thus increase inter-province travel between Ouagadougou and other provinces.

Upon completion of the mathematical model, calibration was preformed using survey data from Marshall et al. (2016), which includes sampling of travelers from sites in Ouagadougou, Saponé, and Boussé during the rainy season of July 2011. The survey by Marshall et al. (2016) is limited since it does not capture movement in other regions such as Houet, located in south-east Burkina Faso; however, data regarding movement and migration in Burkina Faso is generally limited. With the limitations of the data set in mind, parameterization of the movement equations was fit by first selecting the value for $\log_e(\rho)$ that offered the best fit. To do so, a "synthetic survey" was performed in MATLAB which iterated through values 0.05 to 1.8 by steps of 0.05 for $\log_e(\rho)$ with a total of 1000 trials for each $\log_e(\rho)$ value, and $\log_e(\rho) = 0.20$ was selected since it offered the most IQR matches as along with a low mean squared error.[2] Next, the fully parameterized model, using the aforementioned $\log_e(\rho)$ and inferred values $\alpha = 1.27$ and $\tau = 1.342$, was run in the simulation and the resulting frequency of trips and the traveled distance to survey data from Marshall et al (2016).

The final step for calibration and validation was ensuring that the appropriate number of interprovincial trips took place. This was conducted by running the simulation 12 months with a population size of approximately 19 million individuals. All travel between all pairs of cells was logged and cell-to-cell travel numbers were scaled up to the province level to allow for comparison to the

---

[2] The source code can be found in the project repository under `/Analysis/Movement`

Marshall et al (2016) survey data and heatmaps of movement were also generated. The survey by Marshall et al (2018) indicates that about 29.1% of the population travels on a yearly basis with a mean number of 3.42 trips. Approximation of the survey results was achieved with a daily circulation rate of 0.00336. This results in an approximately 19% of all trips to cells within the same province while the remainder leave the province. Ultimately the movement model and its parameterization results in individual movement within the model that is consistent with movement focused on major population centers and follows national transportation networks.

### 5 Immune Response

#### *5.1 Sporozoite Challenge*

To improve the realism of the transmission of the parasite, the model was updated so that individuals would undergo a sporozoite challenge when selected for infection. The challenge was modeled with the presumption that the likelihood of infection decreases as the individual is repeatedly exposed to the parasite, but there remains a small probability of infection, and that a bite by an infectious mosquito does not necessary ensure infection (Churcher et al. 2017).

$$Pr_{inf} = \begin{cases} Pr & if\ \theta < 0.2 \\ 0.1 & if\ \theta > 0.8 \\ Pr\left(1 - \frac{\theta - 0.2}{0.6}\right) + 0.1\left(\frac{\theta - 0.2}{0.6}\right) & otherwise \end{cases} \qquad \textbf{Eq. 5}$$

To allow for these possibilities, Eq. 5 was developed where *Pr* is the probability of infection in an immunologically naïve individual, $\Theta$ is the individual's current immunity, and $Pr_{inf}$ is the final probably of infection. The individual is then considered to be infected by the model if the result of a uniform random draw is less than $Pri_{nf}$.

The validity of the challenge was tested by running the simulation to equilibrium with 100,000 individuals using previously published baseline parameters (Nguyen et al. 2015), at which EIR and *Pf*$PR_{2\text{-}10}$ values are collected (Figure S4). The model presumes that all bites inflicted upon individuals are potentially infectious, yielding the EIR value. The configuration allowed for naïve individuals to be infected 55% of the time, which is consistent with the midpoint of the probability of infection between high and low sporozoite loads in mosquitoes (Churcher et al. 2017). Otherwise, the probability of infection is adjusted based upon the individual's immunity per Eq. 5 with the lower bound of infectious probability set at 10%. To allow for a comparison to the reference data, [3] approximately 30% of the infected population seeks treatment and receives a three-day course of DHA-PPQ with perfect compliance.

**Figure S4.** EIR versus *Pf*PR$_{2\text{-}10}$ for the sporozoite challenge validation. Note that while the model results trend lower than the reference data, the points of reference represent various forms of testing (i.e., ELISA and microscopy) and localities in comparison to the idealized model.

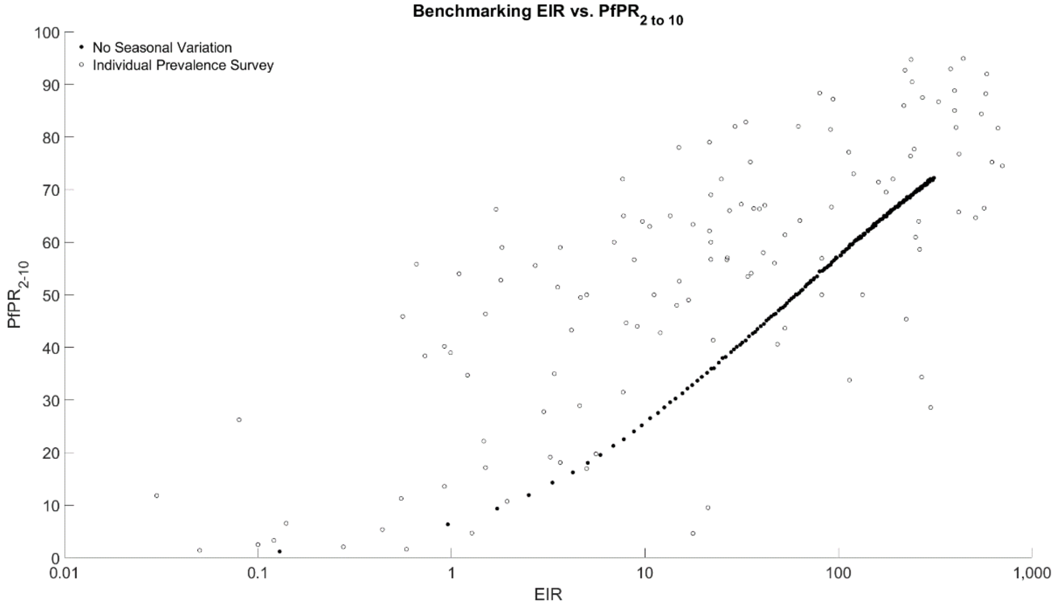

### *5.2 Progression to Symptoms*

Following successful infection with the parasite, the next major process in the immune response is determining if the individual will progress to clinical symptoms or not. The complete process for doing so

[3] Dataset prepared by Malaria Modeling Consortium members (Hay et al. 2005; Cameron et al. 2015) based upon data from: Appawu et al. 2004; Babiker et al. 1997; Barnish et al. 1993; Beier et al. 1990; Beier et al. 1994; Björkman et al. 1985; Bockarie et al. 1994; Bødker et al. 2003; Boudin et al. 1991; Carnevale et al. 1988; Carnevale et al. 1992; Diallo et al. 1998; Diallo et al. 2000; Elissa et al. 2003; Ellman et al. 1998; Gazin et al. 1998; Ijumba et al. 1990; Kabiru 1994; Karch et al. 1993; Koram et al. 2003; Le Goff et al. 1992; Lindsay et al. 1990; Manga et al. 1993; Mbogo et al. 1993; Mbogo et al. 2003; Mendis et al. 2000; Meunier et al. 1999; Mnzava 1995; Nzeyimana et al. 2002; Oloo et al. 1996; Quakyi et al. 2000; Richard et al. 2006; Robert et al. 1986; Robert et al. 1998; Rossi et al. 1986; Seng et al. 1999; Shiff et al. 1995; Shililu et al. 1998; Smith et al. 1993; Sokhna et al. 2004; Thomson et al. 1994; Trape 1987; Trape et al. 1992; and Trape et al. 1994.

is well described in the supplemental materials for Nguyen et al. (2015), although the function describing the probability of progressing to clinical symptoms necessitates examination:

$$Pr_{clin} = \max\left(\frac{0.99}{1 + (\frac{\Theta}{\Theta_{mid}})^{z}}, 0.005\right) \qquad \textbf{Eq. 6}$$

Where $\Theta$ describes the level of immunity, $\Theta_{mid}$ describes the point at which immunity confers a 50% of developing symptoms, and $Z$ is varied between 2.0 and 8.0 to determine its effects upon clinical malaria. To set the lower limit on the probability of developing symptoms, the maximum of $Pr_{clin}$ or 0.005 is used as the final probability of developing clinical symptoms. Since the real value of all variables is unknown, the equation was fit to the data that is available for Burkina Faso (Figure S5). Following simulations with a population of 50,000 individuals, the best fit for under-5 and over-5 estimated clinical cases was found when $M_{mid}$ = 0.15.

**Figure S5.** Curves describing the probability of progressing to clinical symptoms following infection with the parasite based upon the host immune level, at various settings for the midpoint value. Note the curve for the 0.15 value which is the calibration value used in the simulation.

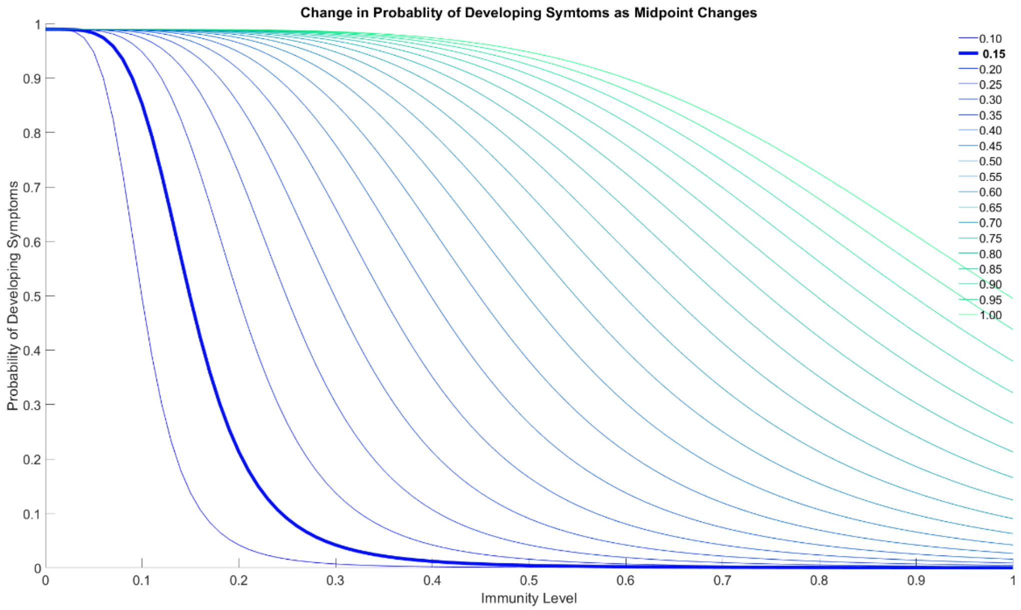

### 6 Treatments

The results of the 2017 – 2018 Malaria Indicators Survey (MIS) was used to inform the treatments that individuals would be given in the model (INSD et al. 2018). This was done by first determining the treatment seeking behavior (i.e., when an individual has symptoms, they attempt to find any treatment for the symptoms) for the under-5 and over-5 population. The under-5 rate is based upon the results of the MIS; however, to account for the discrepancy in treatment seeking behavior in adults (Robyn et al. 2012), the over-5 rate was adjusted to 45% of the under-5 rate (Fig. 1d). The second way that the MIS informed the treatments was via the actual treatment that would be administered. At the time the survey was taken, approximately 79% of children treated for uncomplicated malaria in the two weeks prior to the survey received an artemisinin combination therapy (ACT) (Table S5). As of 2019, an ACT in the form of artemether–lumefantrine (AL), artesunate-amodiaquine (ASAQ), or dihydroartemisinin-piperaquine (DHA-PPQ) are the recommended first line therapy for uncomplicated malaria (President's Malaria Initiative 2019). Although, the use of ASAQ is being phased out in favor of DHA-PPQ due to the expansion of seasonal malaria chemoprevention (SMC).

In preparing the treatments for the simulation some assumptions were made. First, it was presumed that their use would be unform throughout the county, i.e., the location of an individual would not impact which first-line therapy they were more likely to receive. Next, since treatments with ACTs are reported in aggregate, the model inputs were normalized and scaled for the appropriate treatments (i.e., AL, ASAQ, ASMQ, and DP). Finally, ASAQ was removed as a first-line therapy due to the phaseout with the current treatment rate being proportionally reallocated to AL and DHA-PPQ as first-line therapies (see Supplemental Table 1 for treatment configurations used in studies).

**Table S5.** Treatments in use in Burkina Faso and the percentage of children with a fever who received the treatment in the two weeks prior to the EIPBF survey (INSD et al. 2018, p.55). Model inputs indicate the adjusted distributions that were used in the simulation.

| **Treatment** | | **Abbreviation** | **Percent Treated** | **Model Inputs** |
|---|---|---|---|---|
| ACT | Artemether–lumefantrine | AL | 79.4% | 68.0% |
| | Artesunate–amodiaquine | ASAQ | | 2.5% |
| | Artesunate-mefloquine | ASMQ | | 1.3% |
| | Dihydroartemisinin-piperaquine | DP | | 4.9% |
| Mono Therapy | Amodiaquine | ADQ | 12.8% | 12.4% |
| | Artesunate | AS | 2.3% | 2.2% |
| | Chloroquine | CQ | 1.6% | 1.5% |
| | Quinine | QUIN | 5.8% | 5.6% |
| Sulfadoxine/pyrimethamine | | SP | 1.7% | 1.7% |

### 7 Model Calibration and Validation

Three metrics were selected as the primary points of comparison for the model calibration. First, movement validation was preformed to ensure that monthly movement approximated the data that is available for Burkina Faso, this is covered in in Section 4. The next point of comparison is simulated mean *Pf*PR$_{2\text{-}10}$ compared to the reference *Pf*PR$_{2\text{-}10}$ for each province. Finally, the last the simulated number of clinical cases and reported clinical cases were compared to prior estimates and shared with collaborators to determine agreement confidential clinical data. Following selection of the calibration points, preparation of the simulation proceeded with parameters being formatted and coded (e.g., population, treatments, etc.). Next, work progressed to determining the value for $\beta$ or the generalized transmission parameter for the simulation (Nguyen et al. 2015).

The first step in determining the value for $\beta$ was to bin the population in each cell according to the natural breaks using Jenks natural breaks optimization. This resulted in the population binned using the break values of 1803, 6116, 21151, 44761, 79353, 150277, and 206607. While this prevents a unique $\beta$ being calculated for each cell, testing found that there is minimal difference between the $\beta$ value within the breaks. Following binning of the population, calibration proceeded to a parameter space search for each cell for the $\beta$ that resulted in a simulated annual mean *Pf*PR$_{2\text{-}10}$ that is approximately equivalent to the estimated mean $PfPR_{2-10}$ from the Malaria Atlas Project for nominal year 2017 (Weiss et al. 2019).[4] As a result, each cell has a $\beta$ that was determined based upon the population bin, treatment rate, and climate zone.

The overall fit of the calculated $\beta$ was determined by running the simulation with the parameterization, movement, and treatments though model year 2023 with no interventions, alterations to treatment seeking behavior, or genotype mutations enabled. This has the effect of allowing the model to reach equilibrium and allows for several years of data to be checked against the expected $PfPR_{2-10}$ values. The population weighted mean of the last five years of the simulation (i.e., 2018 – 2023) for each province was then plotted in comparison to the expected values to verify the fit (Figure S6). As the national level data showed good agreement with expected values, the cellular data was also checked to ensure consistency. Generally, it appears that cells with low populations (i.e., < 1803 individuals) tend to exhibit more stochasticity than cells with high population. As a result, slightly more variance is anticipated when model outputs are plotted spatially (i.e., genotype frequency heatmaps) although aggregation of results to the province, distinct, or national level corrects for this.

---

[4] While an updated data set (version 2020) has been released by the Malaria Atlas Project offering support up to nominal year 2019, the original $PfPR_{2-10}$ values were used due to concerns that the inclusion of the MIS 2017 survey in the version 2020 results are biased towards lower-than-expected values due to the survey being conducted during the dry season.

**Figure S6.** Verification metrics for the simulation, as reproduced from Figures 1a and 1b of the manuscript. (Left) Note that the population weighted mean *Pf*PR2-10 values are in close agreement with the reference values from the Malaria Atlas Project (Weiss et al. 2019). (Right) Total clinical cases projected at a provincial level are in good agreement with the national incidence rate of 605 (95% credible interval 360 to 990) per 1000 predicted by Rouamba et al (2020). However, as expected, the total reported clinical cases per 1000 can be significantly lower on a provincial level.

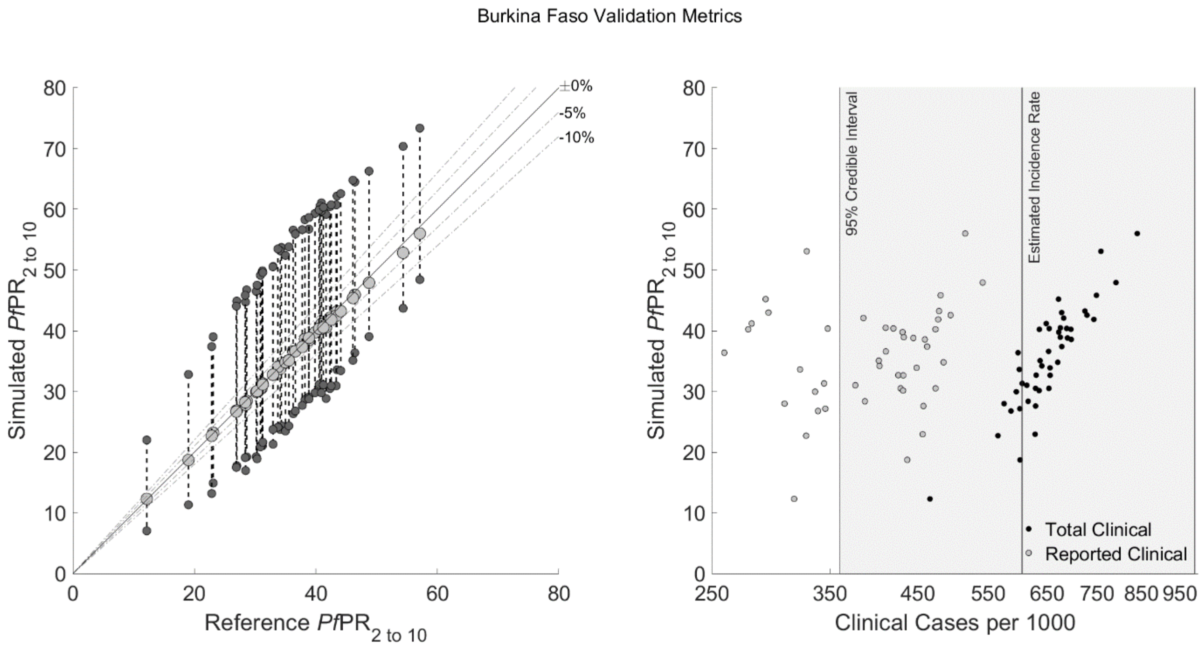

### 7.1 *Statistical Analysis of Policy Options*

The statistical significance of the drug policy options in relation to the status quo scenario were evaluated using the Wilcoxon Rank Sum Test. A MATLAB script was written that selected the maximum 580Y for the last year of each replicate of the relevant policy option and status quo scenario.[5] The same process was used to compare the treatment failure rate. As demonstrated by Table S6, the drug policy options resulted in statistically significant outcomes in comparison to the status quo scenario.

[5] Found in the project repository under `/Source/Figures/bfa_ranksum.m`

**Table S6.** Statistical significance of drug policy options versus the status quo, as computed using the Wilcoxon Rank Sum Test.

| Policy Option | Significance (versus Status Quo) | |
|---|---|---|
| | **580Y Frequency** | **Treatment Failures** |
| Rapid Elimination | $p < 0.0001$ | $p < 0.0001$ |
| Rapid Elimination, Rapid MFT | $p < 0.0001$ | $p < 0.0001$ |
| Rapid Elimination, 10 Year MFT | $p < 0.0001$ | $p < 0.0001$ |
| 10 Year Elimination | $p < 0.0001$ | $p < 0.0001$ |
| 10 Year Elimination, Rapid MFT | $p < 0.0001$ | $p < 0.0001$ |
| 10 Year Elimination, 10 Year MFT | $p < 0.0001$ | $p < 0.0001$ |
| MFT (60% AL, 40% DHA-PPQ) | $p < 0.0001$ | $p < 0.0001$ |
| MFT (70% AL, 30% DHA-PPQ) | $p < 0.0001$ | $p < 0.0001$ |
| MFT (80% AL, 20% DHA-PPQ) | $p < 0.0001$ | $p < 0.0001$ |
| MFT (90% AL, 10% DHA-PPQ) | $p < 0.0001$ | $p < 0.0001$ |
| AL Only | $p < 0.0001$ | $p = 0.0276$ |

### 8 Sensitivity to Mutation Rate

In order to assess the sensitivity to of the model to changes in the mutation rate additional scenarios were performed in which the mutation rate was altered to be very fast (0.01983), fast (0.009915), slow (0.0003966), and very slow (0.0001983) in relation to the calibrated mutation rate (0.001983). The status quo scenario along with the rapid private market elimination and balanced AL/DHA-PPQ multiple first-line therapies (MFT) where adjusted to use the relevant mutation rate and fifty replicates were run for each scenario. The statistical significance of outcomes (Table S7) was computed using the same process outlined previous (see Section 7.1) with the results of the new mutation rate being compared about the comparable scenario with the calibrated mutation rate (e.g., the rapid private market elimination with a very fast mutation rate was compared to rapid private market elimination with the calibrated mutation rate).

Figure S7 through Figure S10 show the results of the alterations to the mutation rate under the status quo scenario with a consistent y-axis. As expected, increases to the mutation rate (Figure S7 and Figure S8) result in an acceleration of the fixation of drug resistant genotypes. In contrast to the baseline scenario the frequency of the KNF genotype is significantly higher than TNF, although this is attributable to the selective pressure of AL usage under the status quo scenario. Additionally, as the mutation rate is slowed (Figure S9 and Figure S10) the time for the 580Y frequency is greatly slowed, although the frequency of 0.01 is reached indicating that fixation is more likely to occur over time. In contrast to the faster mutation rates, the slow mutation rate also results in more variance between replicates and the emergence of the KNF genotype is delayed under the slowest mutation rate.

**Figure S7.** Model behavior under a modified status quo scenario with a very fast mutation rate (10x faster than baseline).

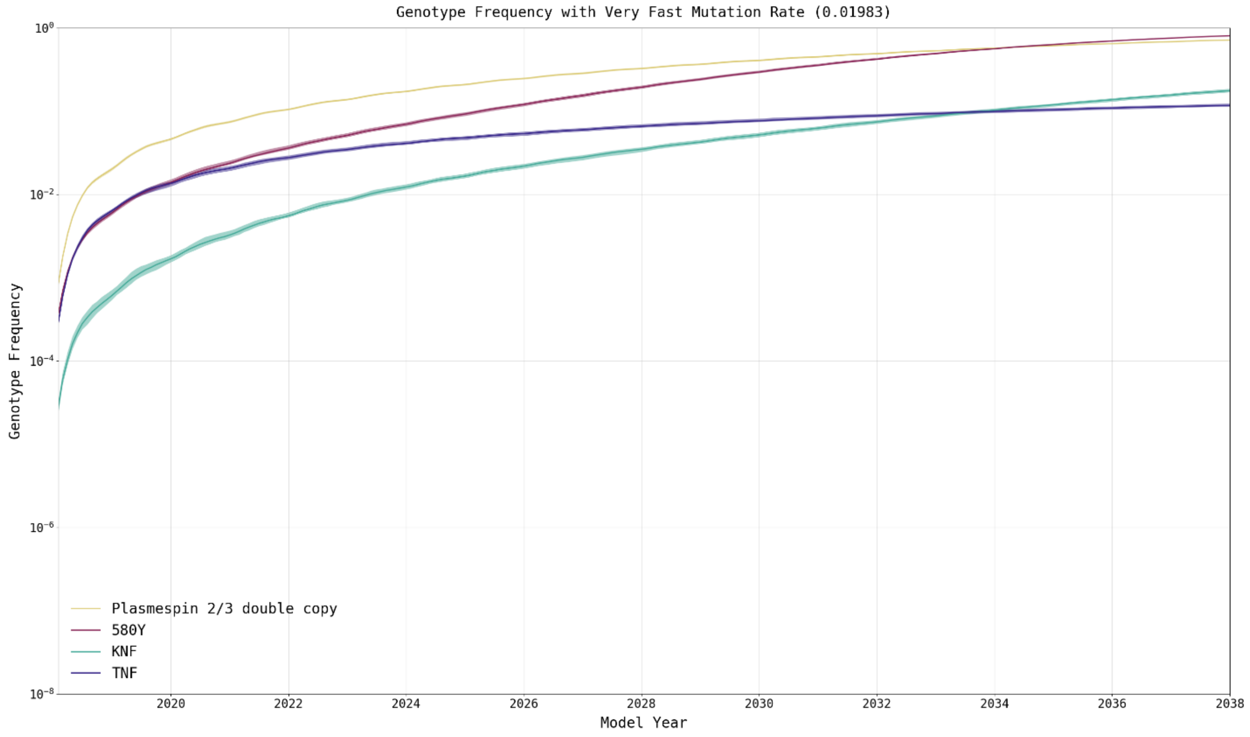

**Figure S8.** Model behavior under a modified status quo scenario with a fast mutation rate (5x faster than baseline).

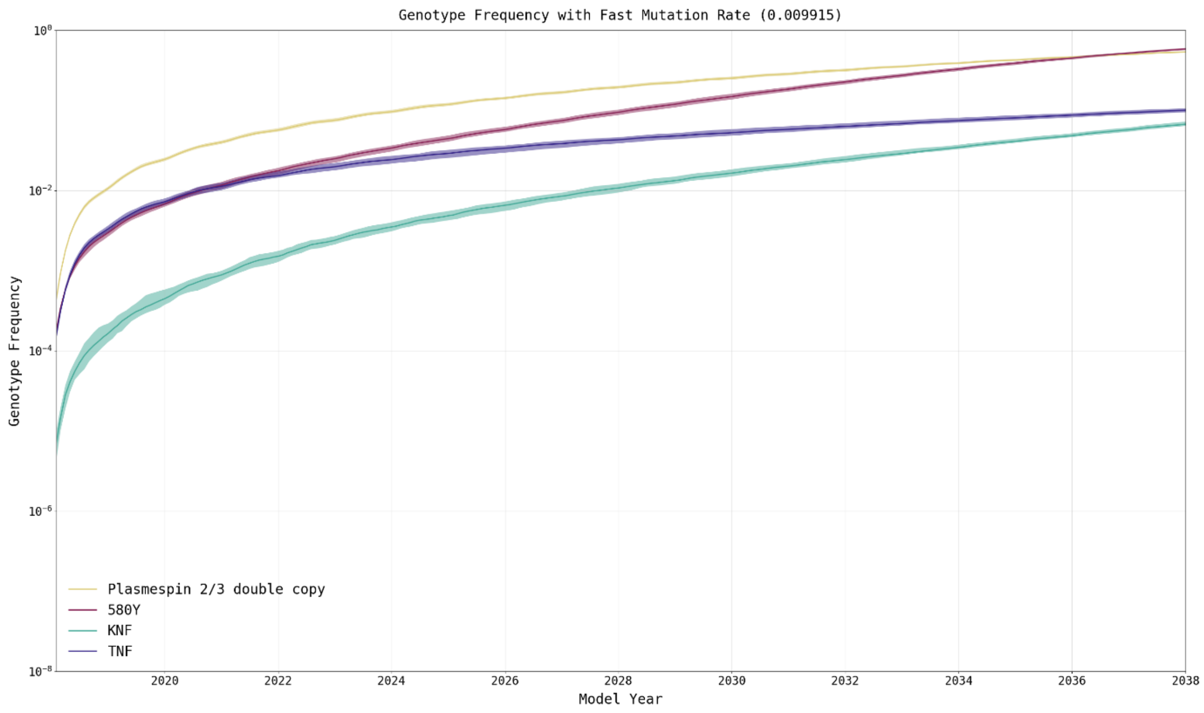

**Figure S9.** Model behavior under a modified status quo scenario with a slow mutation rate (5x slower than baseline).

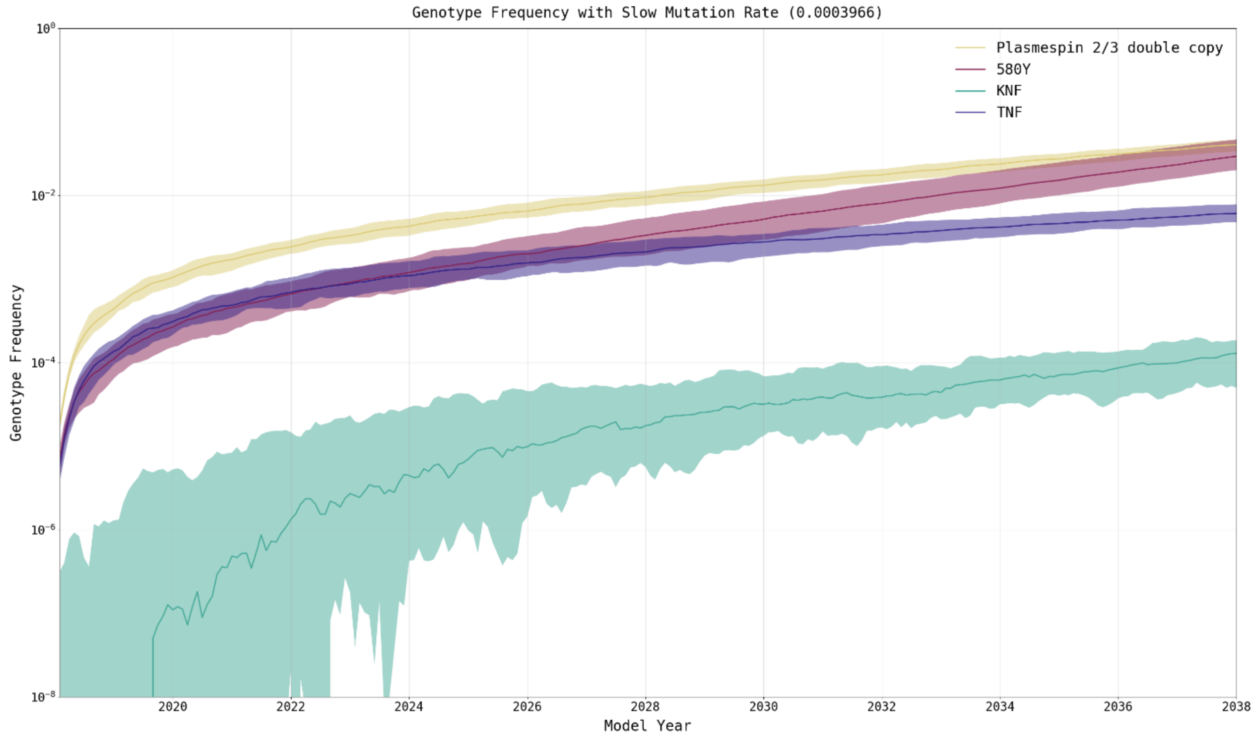

**Figure S10.** Model behavior under a modified status quo scenario with a very slow mutation rate (10x slower than baseline).

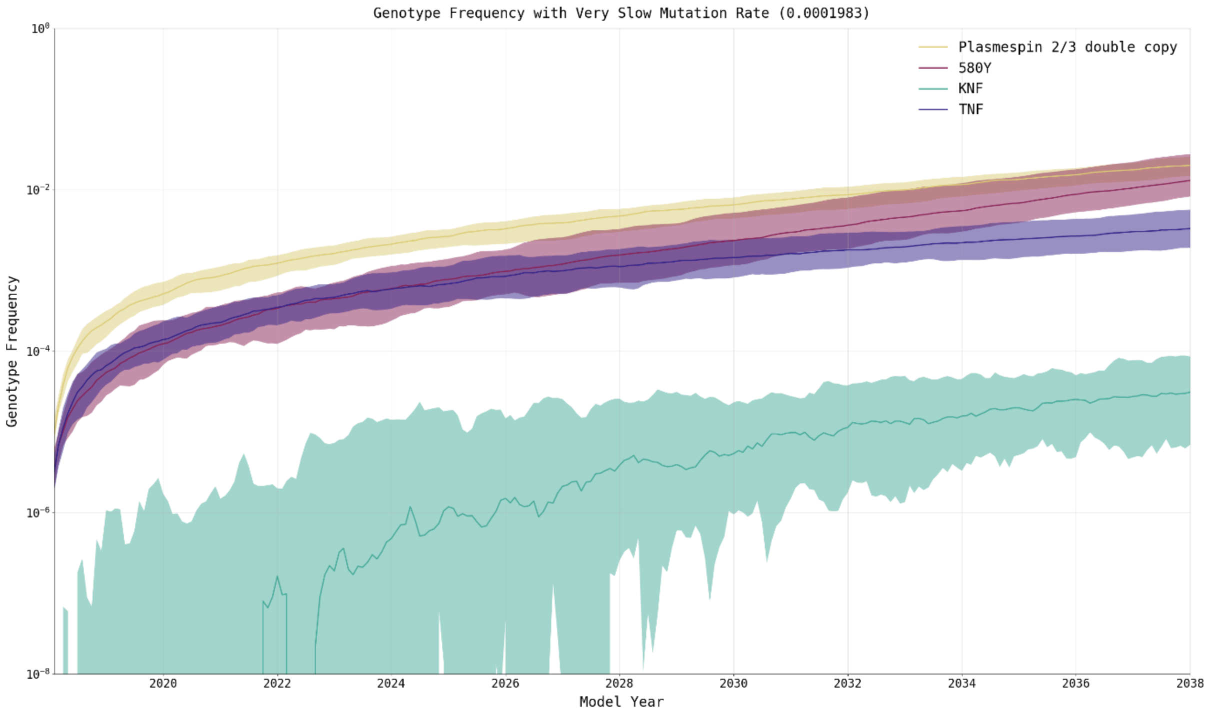

**Table S7.** Statistical significance of changes to the mutation rate versus their baseline mutation rate, as computed using the Wilcoxon Rank Sum Test.

| **Scenario** | **Significance (versus Baseline Mutation Rate)** | |
|---|---|---|
| | **580Y Frequency** | **Treatment Failures** |
| Status Quo, Very Fast | $p < 0.0001$ | $p < 0.0001$ |
| Status Quo, Fast | $p < 0.0001$ | $p < 0.0001$ |
| Status Quo, Slow | $p < 0.0001$ | $p < 0.0001$ |
| Status Quo, Very Slow | $p < 0.0001$ | $p < 0.0001$ |
| Rapid Elimination, Very Fast | $p < 0.0001$ | $p < 0.0001$ |
| Rapid Elimination, Fast | $p < 0.0001$ | $p < 0.0001$ |
| Rapid Elimination, Slow | $p < 0.0001$ | $p < 0.0001$ |
| Rapid Elimination, Very Slow | $p < 0.0001$ | $p < 0.0001$ |
| AL/DHA-PPQ MFT, Very Fast | $p < 0.0001$ | $p < 0.0001$ |
| AL/DHA-PPQ MFT, Fast | $p < 0.0001$ | $p < 0.0001$ |
| AL/DHA-PPQ MFT, Slow | $p < 0.0001$ | $p < 0.0001$ |
| AL/DHA-PPQ MFT, Very Slow | $p < 0.0001$ | $p < 0.0001$ |

### 9 Seasonality in Genotype Frequency

One unexpected pattern observed in simulation output is a seasonal variation in the genotype frequency for drug resistance markers such as the 580Y genotype. In order to eliminate the possibility of these being due to a model artifact additional investigation was undertaken. Following elimination of errors in the source code or data sets used in the simulation, a working hypothesis was developed that the seasonality was correlated to the population immunity to the infection by the parasite.

In order to test this hypothesis smaller simulations were conducted using a single 25 sq.km cell, a population 10,000 individuals, and the general parameterization for Burkina Faso. The use of a smaller simulated space allowed for the capture of the mean of all individual immunity levels, referred to as *theta,* where the individual immunity is described by the variable *M* (Section 5.2). Plotting *theta* revelated clear seasonal variation to the immunity. The seasonal variation becomes more apparent when seasonal transmission is shortened and there is a significant difference transmission intensity (Figure S11). When seasonality is disabled in the simulation and transmission is allowed to persist throughout the year the fluctuations in *theta* disappear (barring typical stochastic variance) and drug resistance genotypes also exhibit a consistent increase in frequency (Figure S12). This further supports the hypothesis that seasonal transmission is driving the seasonal variation and can be confirmed by artificially switching a highly seasonal environment to one in which transmission is persistent throughout the year (Figure S13).

The results of the investigation suggest that the seasonal decline in drug resistance frequency during the dry season appears to be an emergent phenomenon resulting from interactions between individual immune response, vector activity, and likelihood of treatment with an ACT. During the rainy season there is more transmission intensity and more immune naive individuals are infected, leading to more symptomatic cases and treatment with an ACT. This applies selective pressure on C580 favoring the 580Y mutation and the transmission intensity also reduces the generation time which allowing more 580Y mutants to potentially be spread. However, during the dry season the transmission intensity declines, leading to fewer infections. Since biting behavior by the vectors in the simulation is weighted to some individuals being bitten more than others (Nguyen et al. 2015), those individuals are more likely to retain and build an immune response, reducing the likelihood of symptomatic cases and treatment with an ACT. Carrying the 580Y mutation is associated with a higher fitness cost compared to C580 (Babiker et al. 2015; Nguyen et al. 2015), so C580 carriers are selected for when the selective pressure of ACTs is reduced. However, as the rainy season returns, the selective pressure is again increased due to expanded treatment with ACTs favoring the 580Y mutation once again. As such, the patterns of seasonal decline in drug resistant genotypes are consent with Babiker et al (2013) who suggested that the higher fitness costs associated with drug resistance may favor drug sensitive parasites when the additional selective pressure high treatment coverage is lifted.

**Figure S11.** Investigation of seasonal fluctuations in 580Y frequency. While not as apparent with the weighted frequency, the unweighted frequency reveals seasonal variation (Top) along with clear seasonal variation in the mean population immunity level, *theta* (Bottom).

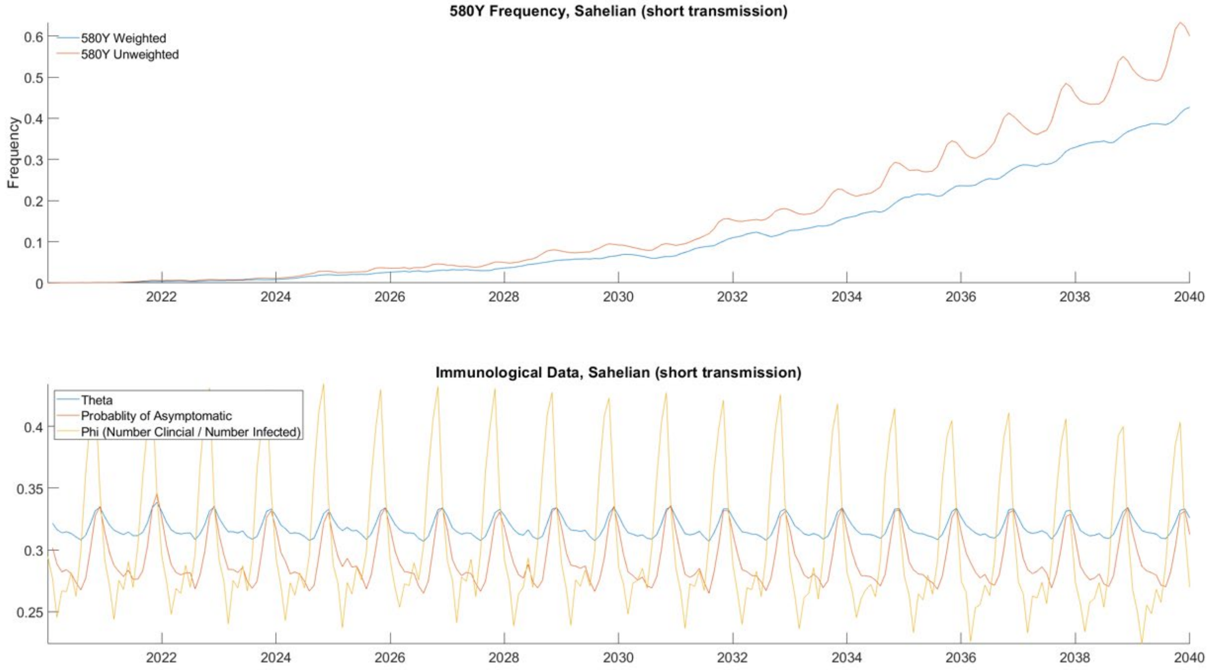

**Figure S12.** Disabling seasonality in the simulation eliminates cyclic behavior with only stochastic variance being observable.

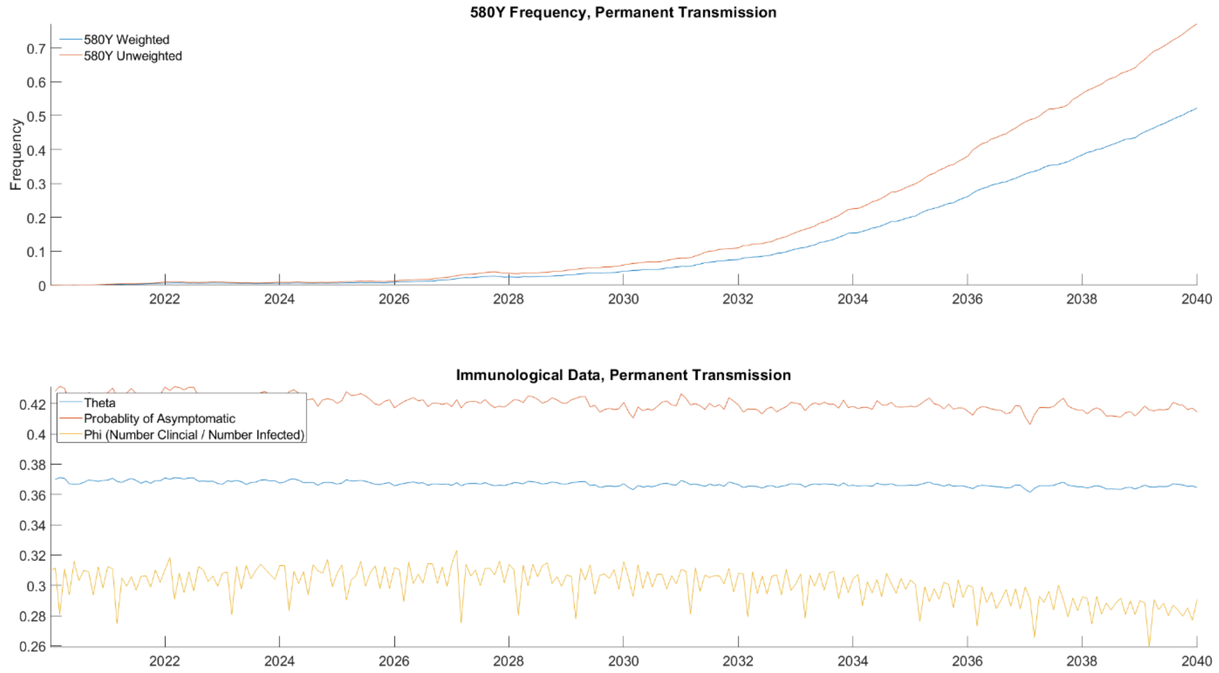

**Figure S13.** Artificially exaggerating the season, along reinforces seasonal behavior, and it can be eliminated by disabling seasonal transmission in favor of persistent transmission in model year 2036.

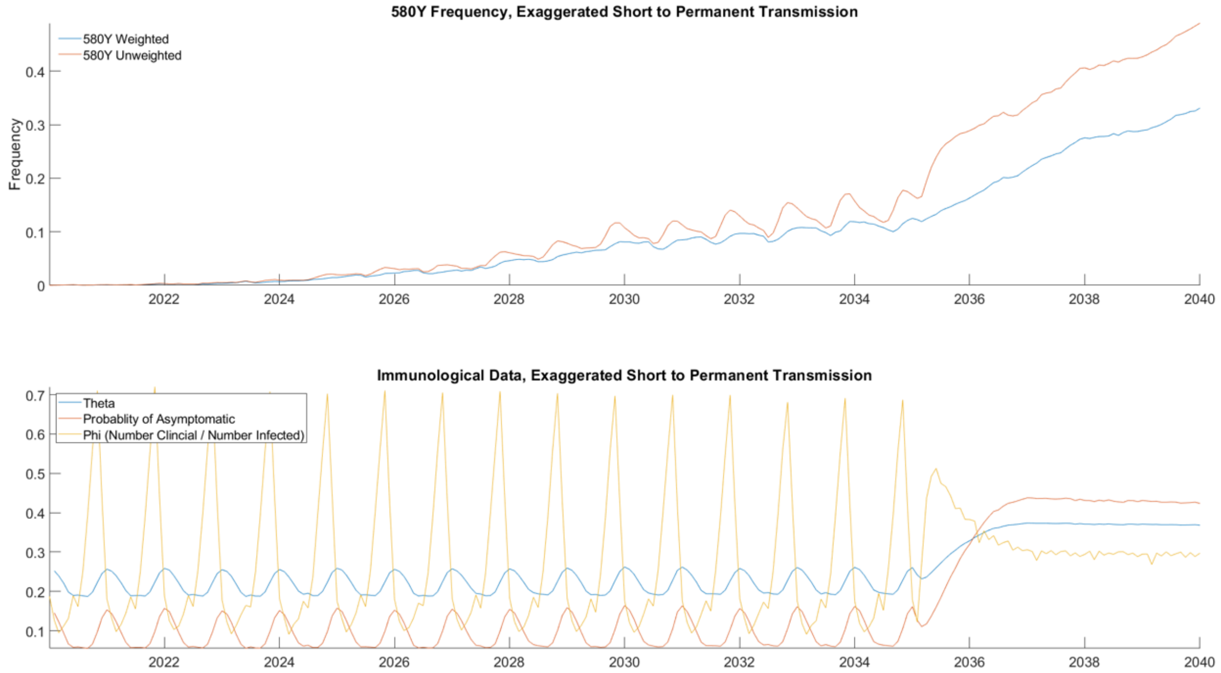
